# Structural variation landscape of Middle Eastern and North African individuals from long-read nanopore sequencing reveals medically relevant variants

**DOI:** 10.64898/2026.02.20.26346743

**Authors:** Talal Al-Yazeedi, Sophie Tandonnet, Sven Hauns, Philip Davies, Sumaya Almansoori, Rolf Backofen, Ahmad Abou Tayoun, Omer S. Alkhnbashi

## Abstract

Structural variants (SVs) are a major source of genomic diversity and disease susceptibility; however, populations from the Middle East and North Africa (MENA) region remain critically underrepresented in global reference databases. We provide a detailed catalogue of structural variation in 61 individuals from diverse MENA countries (8 countries), using publicly available ultra-long Oxford Nanopore sequencing. A scalable and dual-reference alignment-based method (GRCh38 and T2T-CHM13) was employed to comprehensively detect and characterise SVs in these samples. A robust multi-caller approach was applied to classify SVs as high-confidence true positives, requiring consensus from at least three callers. This approach identified 97,765 SVs using GRCh38 affecting 11.6 Mb and 176,494 SVs against T2T-CHM13 affecting 12.2 Mb, the latter providing improved alignment and variant detection. Significantly, 20.3% of the GRCh38 SVs identified in the MENA population were previously unreported, highlighting the region’s uncharted genomic diversity. We identified population-specific structural variants within genes linked to diseases, pharmacogenetics, and immune responses, including SVs nearly fixed within MENA individuals overlapping with OMIM (Online Mendelian Inheritance in Man database) exons. Additionally, by integrating data from the 1K- ONT SV catalogue, alongside chimpanzee and archaic hominin genomes, shared ancient variants were identified, and MENA-specific SVs were highlighted. Evaluating the clinical utility of our resource for patients with MENA ancestry illustrated that it reduces the interpretation burden of SVs in these patients by 92.7% in combination with the 1kGP catalogue (and up to 97.5% with HGSVC2 and 1kGP catalogue combined). These findings establish a foundational reference for structural variants relevant to MENA populations.

## Introduction

Structural variations (SVs), including insertions, deletions, duplications, inversions, and translocations, represent an important aspect of human genetic diversity and have a substantial influence on phenotypic traits, gene regulation, and disease susceptibility. SVs are defined as genomic rearrangements larger than 50 base pairs (bp) and, in total, affect more nucleotides per genome than single-nucleotide polymorphisms (SNPs). However, their biological importance has often been underestimated, mainly due to technological constraints (Audano et al. 2019; Sudmant et al. 2015a). The development of SV detection algorithms for short-read sequencing (SRS) technologies improved our understanding of SVs and their functional implications in diverse global populations. However, traditional short-read sequencing (SRS) technologies, while effective at detecting point mutations and small insertions or deletions (indels), are not well-suited to identify large and complex structural rearrangements, especially those hidden within repetitive and complex regions of the genome (Ebert et al. 2021; Chaisson et al. 2019; Collins et al. 2020; Abel et al. 2020; 1000 Genomes Project Consortium et al. 2012; Almarri et al. 2020). The advent of long-read sequencing (LRS) technologies, particularly those developed by Oxford Nanopore Technologies (ONT), has significantly improved our ability to characterise structural variants (SVs) at base-pair resolution(Schloissnig et al. 2025; Gustafson et al. 2024). These platforms generate read lengths ranging from tens to hundreds of kilobases, enabling better mapping of repetitive regions and more accurate detection of insertions, deletions, and complex rearrangements. As a result, LRS technology has enabled the identification of previously inaccessible SVs and comprehensive SV analysis at the population level.

Recently, numerous studies have made significant strides using LRS technologies, combining ONT with other technologies to construct assemblies for diverse human populations(Logsdon et al. 2025; Kim et al. 2025; Littlefield et al. 2024; Kulmanov et al. 2025; Liao et al. 2023a; Gao et al. 2023; Ghorbani et al. 2025). Notably, the Human Pangenome Consortium (HPRC) has recently released a draft pangenome from 44 diploid long-read sequencing runs and illustrated how this graph reference enhances SV detection(Liao et al. 2023a). Moreover, the Human Genome Structural Variation Consortium (HGSVC) produced near-complete assemblies for 65 diverse human genomes incorporating different technologies and assembly algorithms. Combining these assemblies with the HPRC assemblies in a pangenome graph

significantly enhanced SV detection from short-read sequence data(Ghorbani et al. 2025). While whole genome assemblies are the most efficient resources to identify complex structure variations, especially in regions not represented in the reference genome, their implementation to achieve gapless telomere-to-telomere assembly at a population scale remains limited, due to demanding computational requirements to run the assembly algorithm and the cost of needing to sequence the same sample using more than one technology to achieve a phased near-complete assembly. To address this gap, Oxford Nanopore Technologies (ONT) LRS were applied to analyse SVs from the 1000 Genome Project samples (1kGP), producing one of the most comprehensive references of structural variations. Nevertheless, comprehensive global representation remains limited (Schloissnig et al. 2025). Therefore, population-specific initiatives addressed the lack of genomic representation in global LRS studies through large-scale ONT sequencing. Notably, ONT long-read sequencing of 945 Han Chinese individuals identified structural variants associated with phenotypic diversity and disease susceptibility, and analysis of 3,622 Icelanders provided insights into the role of structural variants in human diseases and other traits (Beyter et al. 2021a; Gong et al. 2025a).

Despite notable progress in the field, structural variants (SVs) remain poorly characterised across many regions worldwide. Most publicly available genomic resources, such as gnomAD-SV, dbVar, and HGSVC, show a marked bias towards individuals of European and East Asian ancestry(Collins et al. 2020; Lappalainen et al. 2013; Ebert et al. 2021). This lack of diverse representation raises serious concerns: ancestry-specific variants may go unnoticed, the functional consequences of SVs might be misunderstood, and the outcomes of association studies may not be relevant to other demographic groups. The Middle East and North Africa (MENA) region exemplifies this disparity. This area features ancient human migration routes, considerable genetic admixture, and distinctive environmental challenges, further enriching a diverse yet largely unexplored genetic landscape (Almarri et al. 2021). Unfortunately, MENA populations are greatly underrepresented in genome-wide association studies (GWAS), sequencing projects, and reference genome initiatives, which prompts critical questions about the comprehensiveness and relevance of genomic research findings. The underrepresentation of certain populations has notable effects on scientific and medical research. Without population-specific variant databases, researchers and clinicians may miss ancestry-enriched or rare structural variants (SVs) that could have clinical importance. While this is already a problem for SNP analysis, it is even a greater problem for the much larger SV. Therefore, adding MENA genomic data into global structural variation maps is essential not only for promoting fairness but also for improving scientific accuracy and medical diagnostics.

To address this significant gap, we analysed published long-read nanopore sequencing of individuals with MENA ancestry and established a scalable and cost-efficient method to discover SVs at a population scale, leveraging the read size offered by Oxford Nanopore Technology. We implemented a dual-reference multi-tool analysis to reduce false positive calls while maximising sensitivity. We characterised the frequency and size distribution of SVs in individuals with MENA ancestry, mostly Gulf Arabs, with additional samples from the wider MENA countries. Our analysis identified a significant number of previously unreported variants and differentiated SVs in MENA compared to the rest of the world population. Our SV resources for the MENA population thus provide a valuable resource to facilitate disease research, and the understanding of phenotypic diversity and the development of personalised medicine.

## Results

### Dual-reference multi-caller SV discovery from nanopore data revealed enhanced mappability and improved detection using T2T-CHM13

Ultra-long (UL) Oxford Nanopore Technology (ONT) reads were obtained from the Sequence Read Archive (SRA) for a total of 61 MENA individuals, who were previously sequenced in two separate genome assembly studies (Nassir et al. 2025; Kulmanov et al. 2025) (Details in method). Those individuals were from diverse Middle Eastern and North African countries, including the United Arab Emirates (42 individuals), Saudi Arabia (9), the Sultanate of Oman (2), Yemen (1), Syria (1), Jordan (1), Egypt (2), and Morocco (2). The obtained ONT raw reads have an average N50 of 56 kb and 147 GB of clean reads per sample (Figure 1A) (Figure S1). The mean length of the longest reads across samples included in this study is 1.4 Mb (Figure S1) (Table S1).

**Figure 1.**
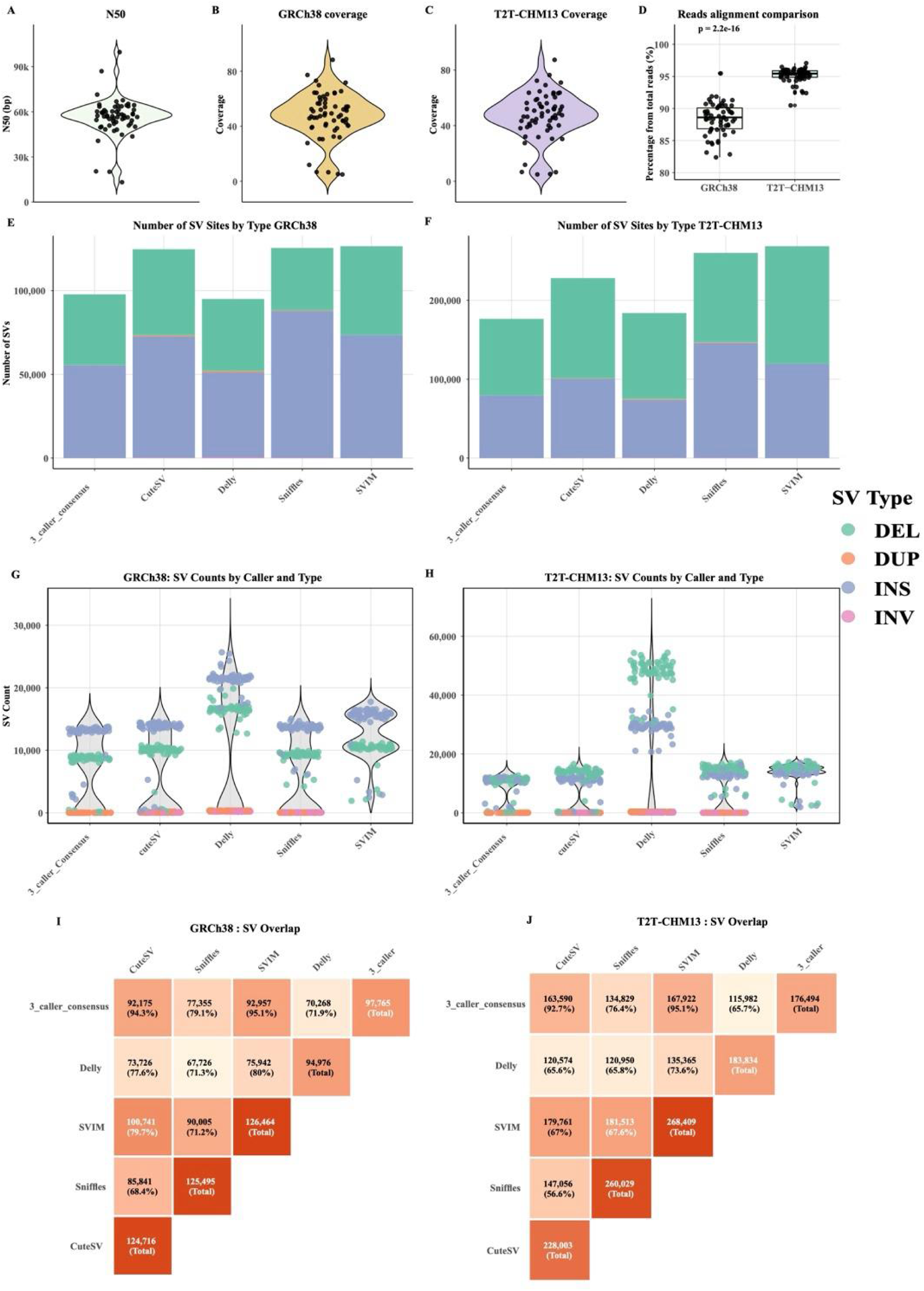
Discovery of MENA SVs from ONT reads using four long-read-based callers and identifying consensus between them. Assessments of read quality for the 61 samples included in this study using a violin plot revealed a mean N50 of 56 Kb (**A**). The complete genome coverage of samples included in this study is represented in violin plots calculated from GRCh38 alignment (**B**) and T2T-CHM13 alignment (**C**). The average coverage across all samples is almost similar in both reference genome alignments, 47.6 for the GRCh38 genome, and 47.1 for T2T-CHM13. (**D**) Boxplot illustrates a significant difference in mean observed when the percentage of reads aligning to GRCh38 compared to CHM13, highlighting a better mapability to the T2T-CHM13 reference. A bar plot for the total number of SVs coloured by type (Deletion in green, duplications in orange, insertions in blue) detected by each caller and in the consensus set, requiring an SV to be called by at least three callers for GRCh38 (**E**) and CHM13 (**F**). Average number of SVs detected per sample, coloured by type, represented in violin plots for each caller and for the consensus sets for the GRCh38 (**G**) and the CHM13 (**H**) reference. Delly showed the most dramatic reference-dependent performance, averaging 77,773 SV calls per sample on T2T-CHM13 versus 38,597 on GRCh38. Concordance between the four SV callers in this study and between each caller and the 3-caller consensus set for GRCh38 (**I**) and CHM13 (**J**).

To understand the impact of genome reference bias and mapping rate, ONT data were aligned to the human genome reference GRCh38(Schneider et al. 2017) and the telomere-to-telomere reference genomes (T2T-CHM13)(Nurk et al. 2022) using parameters tailored specifically for ONT reads(Chen et al. 2024) (Details in methods) (Figure S1, Table S2 and Table S3). The average genome coverage across all samples is similar against both genomes, GRCh38 (47.6x) and T2T-CHM13 (47.1x). (Figure 1 B and C). However, the proportion of reads aligning to T2T-CHM13 (mean 95.2%) was significantly higher than to GRCh38 (mean 88.4%) across samples (paired t-test p < 2.2e-16 and Wilcoxon rank sum test p < 1.13e-11) (Figure 1D).

SVs were discovered using GRCh38 and T2T-CHM13 reference genomes using four long-read-based SV callers: CuteSV(Jiang et al. 2020), Delly (Rausch et al. 2012), Sniffles (Smolka et al. 2024), and SVIM (Heller and Vingron 2019), and then were merged using Jasmine (Kirsche et al. 2023; Gustafson et al. 2024). High-confidence SVs were defined as SVs called by at least three callers. Individual callers identified between 94,976 and 126,464 SV sites for GRCh38 and between 183,834 and 268,409 SV sites for T2T-CHM13, with the latter consistently enabling higher detection across all tools (Figure S2 and S3) (Table S4). The improved detection with the T2T-CHM13 reference genome is due to its gapless nature and to the complete resolution of repetitive and complex genomic regions (Chen et al. 2024; Schmitz et al. 2025; Aganezov et al. 2022). High-confidence SVs requiring at least three callers’ consensus yielded 97,765 (GRCh38) and 176,494 (T2T-CHM13) in total. We detected more high-confidence insertions (56.6%) compared to deletions (43.4%) using the GRCh38 genome reference; however, we observed the opposite using the T2T-CHM13 reference genome (55.1% deletions, 44.9% insertion) (Figure S4), highlighting the difference in SV type detections based on the underlying genome reference (Figure 1 G and H). While inversions were detected by individual callers, the ensemble approach eliminated all inversions, indicating the inherent difference in inversions calling between long-read callers and the downside of our three-caller consensus approach. On average, we identified 20,575 SVs (affecting 11.6 Mbp) and 20,984 SVs (affecting 12.2 Mbp) per individual when using the GRCh38 and the T2T-CHM13 reference genomes, respectively. This is comparable with previous studies discovering SVs using LRS (Gustafson et al. 2024; Audano et al. 2019; Beyter et al. 2021a; Gong et al. 2025a) (Table S6, S7 and S8) (Figure S2) (Supplemental note 1). These calls are approximately four times higher than the average number of SVs called per sample using short-read sequencing in 6,141 Qatari individuals (Aliyev et al. 2026). High-confidence SVs for GRCh38 and T2T-CHM13 were further investigated for allele frequency, size distribution, and genomics consequences in this study.

### Allele frequency and size distribution revealed that large structural variants are the rarest

The site frequency spectrum is skewed under a standard population-genetic model such that the number of distinct variant loci decreases as allele count increases, reflecting the combined effect of mutation, drift and selection (Evans et al. 2007). As expected, both the T2T-CHM13 and GRCh38 high-confidence SVs show a negative correlation between allele count and the number of SVs, in accordance with other studies (Gong et al. 2025a; Beyter et al. 2021a; Schloissnig et al. 2025) and aligning with the allele frequency distribution observed for single-nucleotide polymorphisms in the human genome (International HapMap Consortium et al. 2007) (Figure 2A). This observation is further confirmed by the low percentage of fixed SVs in both genomes (8.4% for GRCh38 and 7.8% for T2T-CHM13) compared to singletons and doubletons (Table S13) (Supplemental note 2).

**Figure 2.**
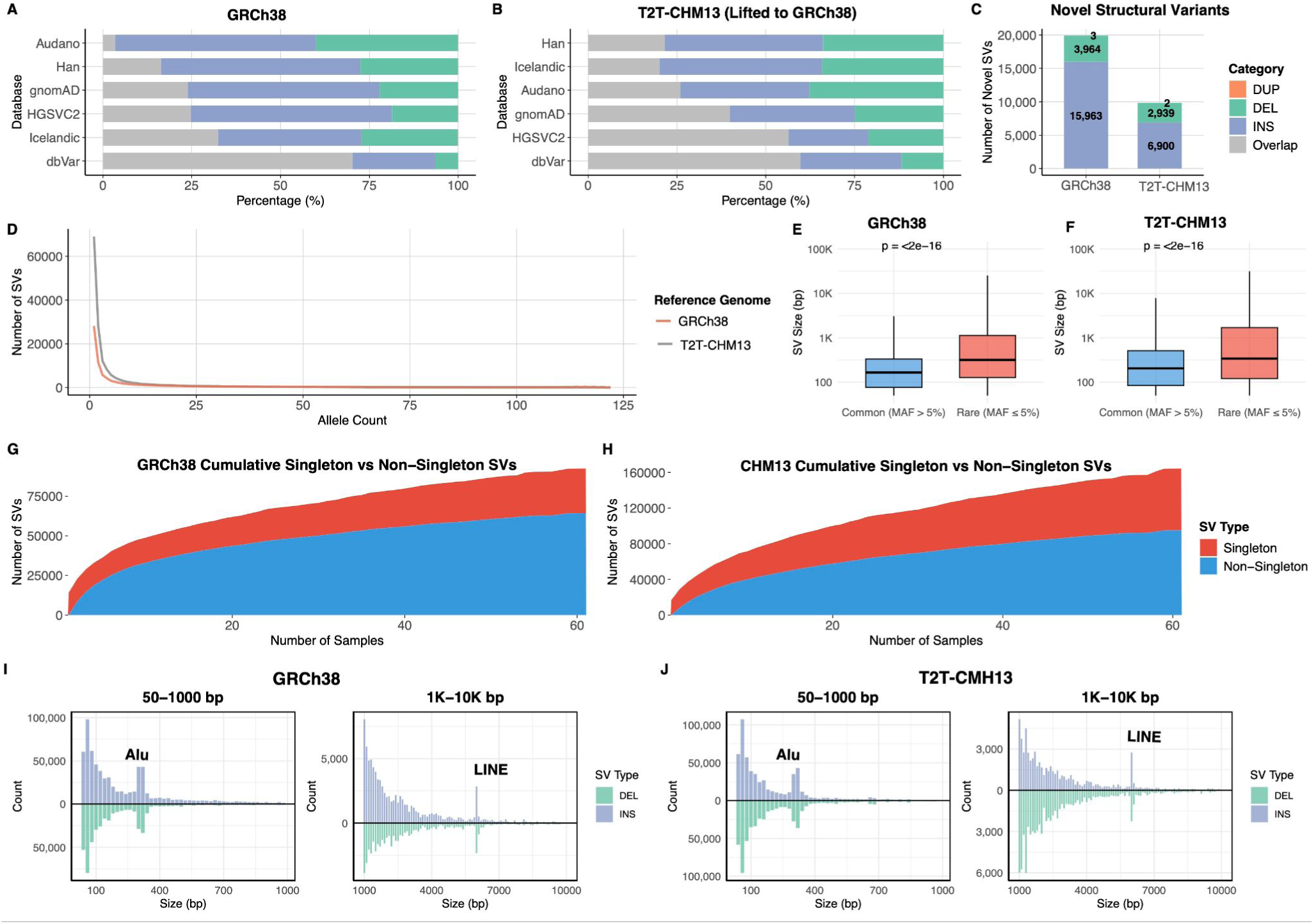
Features of MENA SVs, while a large proportion of MENA ONT-based SVs were undiscovered. Number of reported (Grey) and unreported SVs (coloured by type: Insertion Blue and deletion in Green) according to comparisons with different studies, GRCh38 SVs comparison is shown in (**A**), and CHM13 SVs lifted to GRCh38 are in (**B**). The X- and Y-axes represent the percentage and the datasets used for comparison, respectively. (**C**) Distribution of novel structural variants mapped to GRCh38 assembly and CHM13 assembly. (**D**) Plotting the distribution of allele count (x-axis) against the number of SVs recorded for each allele count (y-axis) reveals a decrease in SV frequency spectrum when SVs become common in MENA individuals (n = 61, allele count = 122). A boxplot comparing SV size distributions between common (MAF > 0.05) and rare (MAF <= 0.05) SVs for GRCh38 (**E**) and T2T-CHM13 (**F**). A cumulative distribution of singleton (Red) versus non-singleton (Blue) in GRCh38 (**G**) and T2T-CHM13 (**H**) when a new sample is added, samples are in the x-axis, and the y-axis represents the absolute count of SVs. The number of singleton SVs quickly becomes common in the population, as the proportion of common SVs increases. Unfortunately, due to the sample size (n=61), we didn’t notice a plateau of the non-singletons curve to determine the number of individuals required to capture the most common SVs in the MENA population. Size distribution of SVs detected against the GRCh38 (**I**) and the T2T-CHM13 (**J**) references. Frequency (y-axis) of SV sizes (x-axis) were separated into two windows for better visualisation, ≥1000 and ≤1000, and insertions (Blue) are separated from deletions (Green). A distinctive peak observed at 300bp represents Alu retrotransposons, and at 6000 represents LINE elements.

Classifying SVs based on Minor Allele Frequency (MAF) revealed that most high-confidence SVs were common, in contrast to what has been observed in other studies, where most discovered SVs were rare (MAF ≤ 5%) (Beyter et al. 2021a; Aliyev et al. 2026; Gong et al. 2025a; Schloissnig et al. 2025). This reflects the small sample size constraint in this study (n=61, 122 alleles, where variants present in just 4 individuals already exceed the 5% MAF), and the UAE-biased cohort (68% of samples). Additionally, the sizes of rare SVs (MAF ≤ 5%) in both T2T-CHM13 and GRCh38 are significantly greater than common SVs (MAF > 5%). The mean size for GRCh38 rare SVs is 1,405 bp, while the mean size for common SVs is 545 bp (paired t-test p < 3.5e-4 and Wilcoxon rank sum test p < 1.38 × 10^-2^). On the other hand, the mean size for CHM13 rare SVs is 1,397 bp, and the mean for common SVs is 728 bp (Wilcoxon rank sum test p < 2.2 × 10^-16^) (Figure 2 B and C) (Table S14). This observation was consistent across deletions and insertions independently and similar to observations in previous studies (Gong et al. 2025a; Schloissnig et al. 2025) (Figure S5). Small SVs (<1 kb) predominated in both datasets (89.1% GRCh38; 82.5% T2T-CHM13), though overall T2T-CHM13 detected a higher proportion of large SVs (≥1 kb) at 17.5% compared to 10.9% in GRCh38 (Table S14).

We observed that the rate of SV discovery increases when new samples were added, singleton and common SVs increase with the addition of a new sample and with no clear sign of a plateau, indicating that the sample size of 61 is insufficient to detect common SVs in MENA individuals (Schloissnig et al. 2025; Gong et al. 2025a) (Figure 2D, E and S5). Furthermore, and in accordance with previous studies, across all callers on both reference genomes, including the high-confidence set, we observed distinct peaks at sizes of approximately 300 and 6000 bps (Audano et al. 2019; Liao et al. 2023b; Beyter et al. 2021b; Gong et al. 2025b; Sudmant et al. 2015b; Gustafson et al. 2024) (Figure 2I and J, S2 and S3). These peaks likely correspond to the retrotransposition activities of Alu and LINE elements, respectively.

### A large proportion of MENA ONT-based SVs were not previously reported

Our investigation revealed a substantial prevalence of previously undocumented structural variants within the MENA population compared with public databases and LRS-based studies using a threshold of reciprocal overlap rate of ≥ 50%, where an SV will be considered reported if the overlapping rate between high-confidence MENA SV and published datasets is ≥ 50%. Comparing our GRCh38 SVs against the gnomAD SVs dataset (Collins et al. 2020) (A high-coverage short-read sequence dataset of 14,891 global samples) identified 74,431 (76.13%) variants that showed no overlap with gnomAD structural variants. Comparison with long-read sequencing of diverse populations, however, with a small sample size of fifteen, such as the Audano et al dataset (Audano et al. 2019), revealed that 96.59% (94,439) of structural variants in our study are not contained in this dataset. Furthermore, assessment against the Human Genome Variation Consortium phase 2 (HGSVC2) (Ebert et al. 2021) dataset revealed 75.22% (73,544) of SVs in our study were not reported. When our SVs dataset is compared to a specific population sequencing using ONT with a large sample size, we detect a remarkably low percentage of overlap, highlighting the unique structural variation of populations from different unrelated ethnicities. Comparison with the SVs discovered from 945 Han individuals identified only 16.36% overlap (15,996 SVs shared). Similarly, comparison with 3,622 Icelanders sequenced using ONTs identified 32.5% overlap (31,736 SVs shared). Finally, even when compared against the comprehensive dbVar repository (Lappalainen et al. 2013), which consolidates findings from 219 distinct studies utilising diverse sequencing technologies, 29.76% (29,095) of our structural variants remained unreported (Figure 2F and Table S15). Overall, 19,930 (20.3%) of SVs from the GRCh38 high-confidence SVs dataset in the present study were not previously documented in any of the investigated publicly available databases. These novel SVs comprise 15,963 insertions, 3,964 deletions, and 3 duplications (Figure 2H). We performed a parallel analysis using our T2T-CHM13 SVs dataset after coordinate liftover to GRCh38 coordinates. This analysis corroborated our initial observations, revealing that 9,841 (11.8%) of lifted T2T-CHM13 SVs to GRCh38 were not detected in publicly available databases (Figure 2J and Table S15).

### MENA Structural variants detected using the T2T-CHM13 are predominantly in repetitive regions

Analysis of SVs overlapping gene regions showed similar numbers of affected genes in GRCh38 (14,815) and T2T-CHM13 (14,138), of which 3,687 and 3,395 genes, respectively, had SVs overlapping at least one exon (Table S17). To examine the intersection with medically relevant variants, we inspected the overlap with the GRCh38 Genome in a Bottle (GiaB) medically relevant genes (n=5,027). We identified 14,506 SVs (14.84% of total SVs) overlapping medically relevant genes in GRCh38, of which 2,127 (2.18% from total SVs) overlap with GiaB classified medically challenging genes. Examination of the intersection with protein-coding regions revealed enhanced overlap using T2T-CHM13, where T2T-CHM13 high-confidence SVs showed 2,328 (2.38%) overlap with coding sequences, while 889 SVs mapped to GRCh38 (0.91%) intersected with RefSeq CDS(Figure 3 and Table S17).

**Figure 3.**
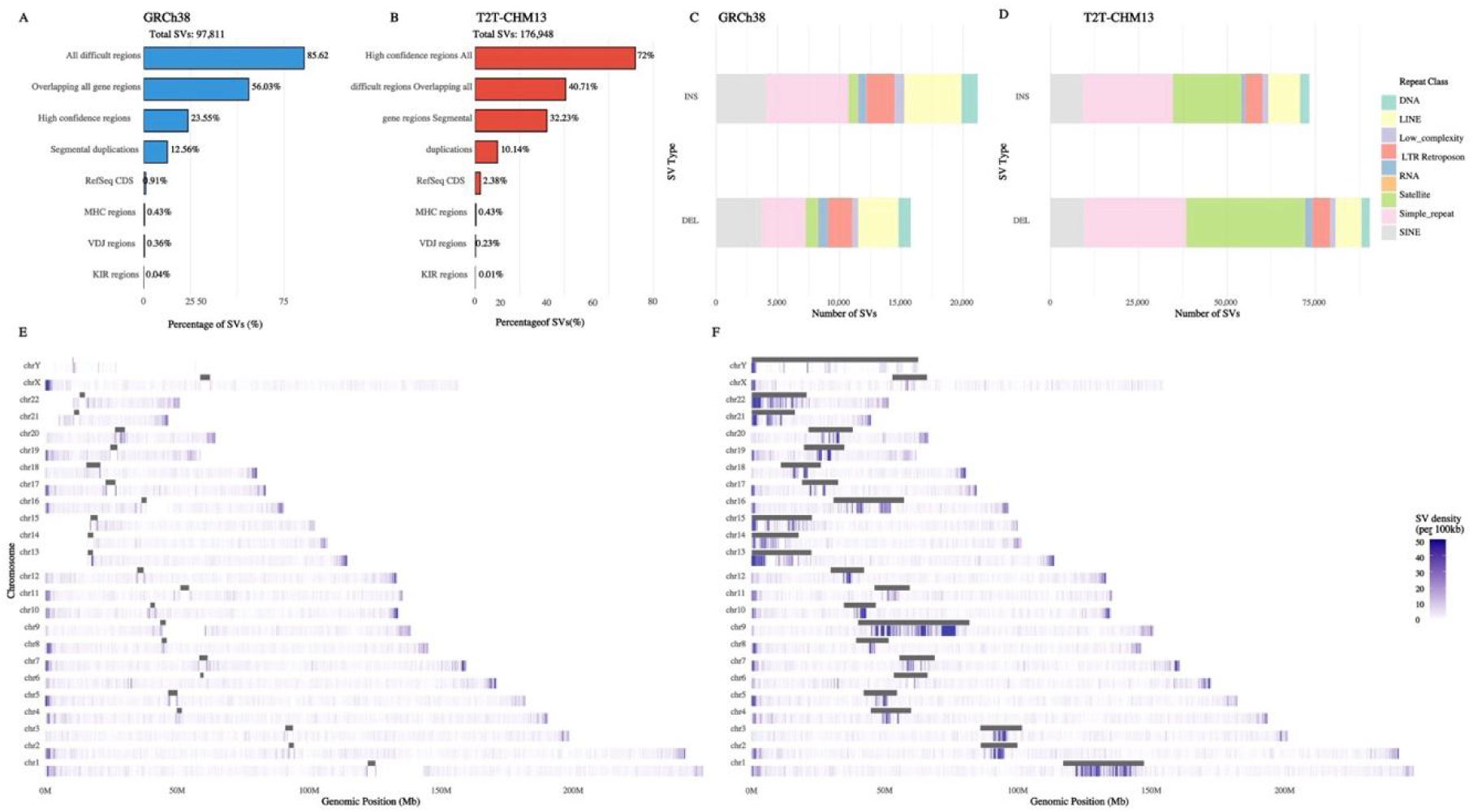
SV distribution in GiaB stratifications and repeats. Percentages of GRCh38 (**A**) and CHM13 (**B**) MENA SVs’ intersection with GiaB stratifications. The majority of GRCh38 SVs (85.62%) were located within all difficult regions compared to T2T-CHM13, where most SVs were in high-confidence regions. GiaB-classified segmental duplication regions intersect with 12.56% of GRCh38 SVs versus 10.14% for T2T-CHM13; this minor difference reflects the better resolution of repetitive sequences in the complete genome assembly. Number of SVs overlapping with repeat classes annotated using RepeatMasker for GRCh38 (**C**) and CHM13 (**D**). SVs density across chromosomes in a 100kb window for GRCh38 (**E**) and CHM13 (**F**). The centromere locations for both genomes are highlighted in a grey bar over the genome track. High density of SVs is observed at the centromeric regions of the CHM13 genome.

Overlapping MENA high-confidence SVs with repetitive elements, where at least one SV breakpoint falls in a repeat region, showed different patterns between the two reference genomes. In the GRCh38 reference, 25.97% of SVs (25,397) overlapped with repeat regions, whereas the T2T-CHM13 reference showed substantially higher overlap, with 77.69% of SVs (137,164) intersecting repetitive sequences. This dramatic difference confirms previous observations and reflects the complete representation of repetitive and previously inaccessible genomic regions in the telomere-to-telomere assembly (Kim et al. 2025; Chen et al. 2024; Audano et al. 2019). Similar to observations from long-read sequencing of samples from the 1kGP, the most prevalent repeat classes overlapping with SVs in both genomes were simple repeats, accounting for 10.47% of GRCh38 SVs and 30.74% of T2T-CHM13 SVs (Schloissnig et al. 2025; Kim et al. 2025). Beyond repeat overlap, we characterised the mechanistic origins and classes of MENA SVs using the Structural Variant Annotator (SVAN) (Schloissnig et al. 2025). SVAN successfully classified 76.0% (74,364) and 77.5% (136,769) of SVs from GRCh38 and T2T-CHM13 datasets, respectively. In accordance with the SVAN annotation of SVs from the 1kGP, variable number tandem repeats (VNTRs) dominated both datasets, 52.0% (50,863) of total GRCh38 SVs and 63.5% (112,004) of total T2T-CHM13 SVs. Duplications comprised 9.5% (9,245) and 8.1% (14,218) of SVs mapped to GRCh38 and T2T-CHM13, respectively, with tandem duplications representing the majority in both references (Figure S8 and S9) (Supplementary note 3).

We assessed the genomic context of our MENA SVs according to the Genome in a Bottle (GiaB) stratification (Dwarshuis et al. 2024). In GRCh38, 85.62% of SVs were located within all difficult regions compared to 40.71% for T2T-CHM13, while high-confidence regions contained 23.55% and 72% of SVs, mapped to GRCh38 and CHM13, respectively (Figure 3 and Table S17). This inverse relationship demonstrates the improved mappability and reduced technical artefacts achieved by the telomere-to-telomere assembly, effectively converting difficult regions into a high-confidence mappable sequence. Within GIAB highly polymorphic and complex immunologically critical regions, SVs were detected in MHC regions, VDJ regions and KIR regions consistently across references that are technically challenging to sequence using short-read (Table S17).

### Integrating MENA SVs with the 1K genome project, ONT SVs identified differentiated and MEAN-specific SVs

We integrated Sniffles-based SVs discovered in MENA with the 1kGP ONT Sniffles SVs (n=1,019) using SURVIVOR (version 1.0.7), to robustly merge SVs called with a similar algorithm. This integration reveals distinct population genetic patterns and reference genome-dependent variant detection (Figure S10, Table S18 and S19).

A Principle component analysis of SVs shared between MENA and 1kGP-ONT individuals, retaining only sites genotyped in ≥ 95% of individuals, revealed that MENA samples clustered distinctly, confirming the MENA distinct SV profile that can not be attributed to a single represented global ancestry (Figure 4A and B). Pearson correlation of SV allele frequencies revealed that MENA populations are most similar to European (r = 0.635) and American (r = 0.624) populations, followed by South Asian (r = 0.613), with lower similarity to East Asian (r = 0.515) and African (r = 0.508) populations. This ranking was consistent using T2T-CHM13 (EUR r = 0.549, AMR r = 0.528, SAS r = 0.502, EAS r = 0.424, AFR r = 0.385) (Supplementary Figure S11 and S12). African populations shared the highest absolute number of SV sites with MENA (GRCh38: 33,247; T2T-CHM13: 71,487), consistent with the larger reservoir of African structural variation, followed by South Asian populations, consistent across both reference genomes (Tables S20), reflecting the Middle East’s role as a corridor for human migration between Africa and Eurasia. Population-specific analysis identified unique MENA GRCh38 SVs, 55 SVs affecting 17 protein-coding genes and 223 T2T-CHM13 SVs affecting 33 protein-coding genes (Figure 4, Table S19).

**Figure 4.**
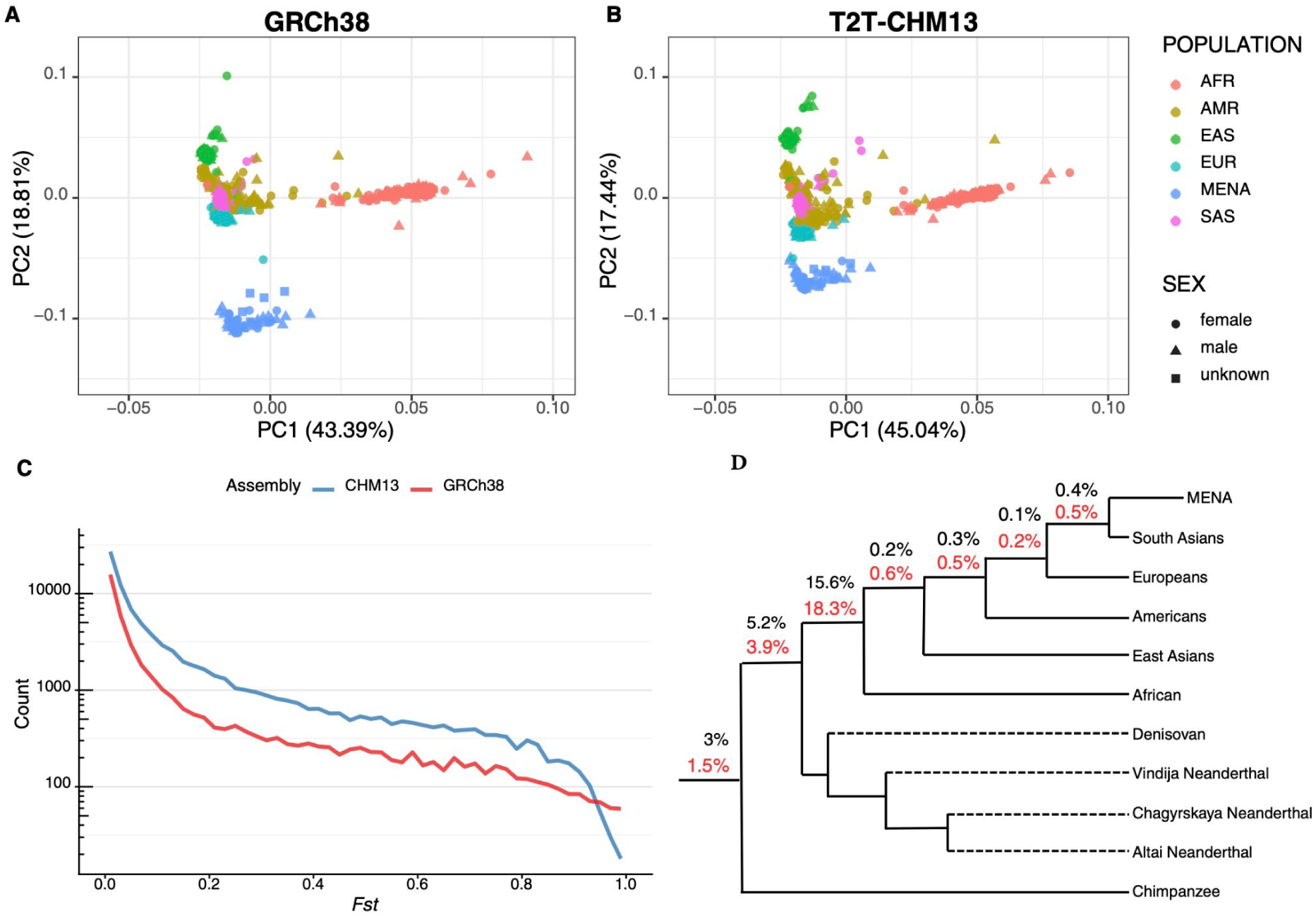
MENA Structural variants merged with global populations. A Principle component analysis of Sniffle-called SVs shared between MENA and 1kGP-ONT individuals, retaining only sites genotyped in ≥ 95% of individuals, revealed that MENA samples clustering distinctly using GRCh38 (**A**) and T2T-CHM13 (**B**) reference genomes. (**C**) Fixation index (*F_st_*) distribution comparing T2T-CHM13 and GRCh38 reference genomes, showing population differentiation patterns. (**D**) Cladogram displaying genetic relationships between global populations and archaic hominins; the percentages indicate the fraction of SVs shared with the MENA population gained at each evolutionary period (GRCh38 in black and T2T-CHM13 in red). Population sharing analysis identified 55 SVs specific to the 61 MENA individuals included in this study using GRCh38 and 223 SVs using CHM13.

To identify differentiated SVs in MENA individuals, the fixation index (*F_st_*) per SV was calculated for the MENA samples against the remaining samples from the 1kGP ONT. Using the GRCh38 reference, we identified 4,068 differentiated SVs (*F_st_* ≥ 0.2), of which 2,260 SVs fall in gene regions affecting 1,202 protein-coding genes and the coding region of 26 genes. Compared to 2,959 differentiated SVs intersecting with 860 protein-coding genes using the T2T-CHM13 reference (Figure 4, Table S21). Cross-reference validation revealed limited variant-level concordance (297 SVs independently differentiated in both assemblies, affecting 133 protein-coding genes), reflecting differences in SV site representation in each assembly and reference-dependent allele frequency statistics. A total of 630 differentiated SVs mapped to GRCh38 intersect with GiaB-medically relevant genes affecting 405 genes, of which 53 genes are GiaB-classified medically challenging genes, highlighting variants in genomically complex areas typically inaccessible to short-read sequencing (Figure S13 and Table S21).

Among the differentiated SVs in GRCh38 GiaB medically relevant genes, two coding sequence variants were identified: a 61 bp deletion in *CEL*, a gene linked to Maturity-onset diabetes of the young type 8 (MODY8) (Sun et al. 2024), was depleted in MENA (*Fst* = 0.222, MENA AF = 0.276, global AF = 0.620–0.655), in addition, a 282 bp insertion in *TRIOBP* (*Fst* = 0.290, MENA AF = 0.442, global AF = 0.540–0.647), a gene associated with autosomal recessive hearing loss (DFNB28), was similarly depleted. At the exonic level, a 316 bp insertion in *HLA-DQB1* (*Fst* = 0.596, MENA AF = 0.271, global AF = 0.003–0.023), encoding the MHC class II beta chain associated with type 1 diabetes and coeliac disease susceptibility, was the only MENA-enriched variant compared to the global population. Four additional exonic variants were depleted in MENA, affecting *CD247* (T-cell receptor signalling), *VPS53* (pontocerebellar hypoplasia), *WDR72* and *KLK4* (amelogenesis imperfecta) (Table S22).

### Functional annotation of the high-confidence MENA SVs revealed transcript deletions and enhanced detection in critical regions using the T2T-CHM13

Functional annotation for the GRCh38 and T2T-CHM13 high-confidence MENA SVs revealed that most annotations were localised to intronic regions, 67.5% of total annotations for GRCh38 and 55.7% for T2T-CHM13. However, SVs in functionally critical categories were also identified, including 2,933 SVs (0.5%) affecting the GRCh38 protein coding sequence (Figure 5A and Table S23). Functional annotation of SVs using the T2T-CHM13 reference showed enhanced detection in critically functional regions, with protein-coding affecting SVs increasing to 0.9% of total annotations (3,575) compared to 0.5% in GRCh38 (Figure 5B and Table S23).

**Figure 5.**
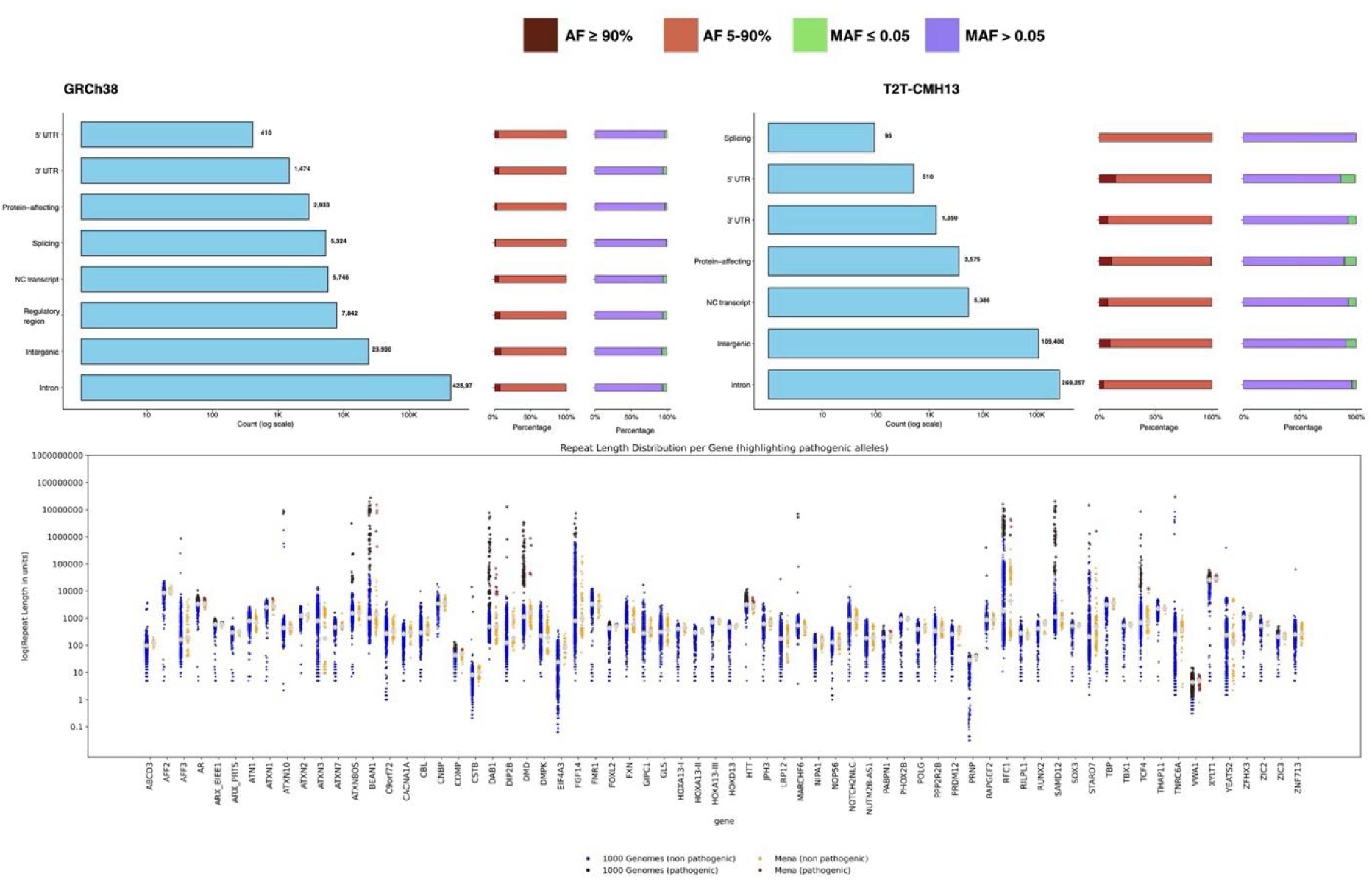
Functional consequences and clinical relevance of MENA structural variants. (**A**) Distribution of variant effect predictor (VEP) annotations for GRCh38 3-caller consensus SVs, showing predominant intronic localisation (67.5%) with functionally critical categories including protein-affecting variants (0.5%), splicing-related variants (0.8%), and non-coding transcript variants (0.9%). (**B**) VEP annotation distribution for T2T-CHM13 3-caller consensus SVs demonstrating enhanced detection of protein-affecting variants (0.9%) and reduced splicing annotations (0.02%) compared to GRCh38, with intronic variants comprising 55.7% of total annotations. (**C**) Comparison of short tandem repeat (STR) lengths between MENA individuals (n=61) and 1000 Genomes Project ONT dataset (n=1,018) across 68 clinically relevant genes. Violin plots show allele-length distributions with notable differences: *EIF4A3* (longer in MENA) and *ATXN3* (shorter in MENA). Red circles indicate pathogenic alleles in MENA, black highlights pathogenic alleles in 1K ONT and grey highlights the mean value.

Amongst the high-confidence SVs in critical categories, we identified a total of 351 unique whole transcript deletion SVs affecting 98 protein-coding transcripts of 52 genes in GRCh38 and 313 unique whole transcript deletion SVs affecting 190 protein-coding transcripts of 130 genes in T2T-CHM13 (Tables S24 and S25). The majority of transcript deletions detected in MENA were observed in at least one population from the 1kGP sniffle-based SVs, indicating these transcript deletions represent shared human variation rather than MENA-specific events. A total of 105 whole transcript deletion SVs in MENA were independently confirmed in both references, through reciprocal coordinate lifting, eight of which affect protein-coding genes (Tables S24). Three SVs, out of the eight, cause whole-gene deletions also detect in the 1kGP ONT sniffles calls: a 5.6 kb deletion harbouring *IGFL3* detected in 3 MENA individuals and present across all 1kGP continental populations (MENA AF = 0.041, global AF = 0.050); a 5.1 kb deletion ablating all six protein-coding transcripts of *TPSAB1* in 1 MENA individual and enriched in East Asian populations (EAS AF = 0.171, global AF = 0.043) located at the structurally complex chr16p13.3 trypaselocus, where copy number variation underlies hereditary alpha tryptasemia (Lyons 2018), and a 8.8 kb deletion encompassing *ZSCAN5C* in three MENA individuals (global AF = 0.016) restricted to African (AFR AF = 0.057) and MENA populations. A whole transcript-deletion SV confirmed in both references affects *HGFAC* (4.2 kb deletion, 2 individuals, MENA AF = 0.018, global AF = 0.001), encoding a hepatocyte growth factor activator. Remaining four cross-reference confirmed transcript-deletion SVs were detected in MENA individuals but absent in 1kGP dataset required further validation in larger cohort including *DZIP1L* (5.4 kb heterozygous deletion, 1 individual), associated with autosomal recessive polycystic kidney disease (OMIM #617610) (Subramanian et al. 2025); *PSG7* (1.5 kb deletion, 3 individuals); and *ATXN3L* and *GRIK4* (16.5 kb and 4.1 kb homozygous deletions, respectively, each detected in a single individual).

### Despite reference discrepancies, dual-reference analysis identifies clinically important structural variants

Reference genome bias was evident when inspecting the functional consequences of SVs at known discrepant regions between GRCh38 and T2T-CHM13 (Gigliotti et al. 2020). At these discrepant loci, including *GSTM1*, *CR1*, *KLRC2*, and *MAP2K3*, SV detection patterns were reference-dependent, with variants appearing as deletions against one reference and insertions against the other, reflecting structural differences between the assemblies (Saitou et al. 2018a; Yang et al. 2023; Gigliotti et al. 2020; Saitou et al. 2018b; Yang et al. 2022; Chen et al. 2024) (Figures S14-S18) (Supplementary note 3). In addition, our dual-reference approach enabled haplotype characterisation at these complex loci. For example, at *KLRC2* is deleted in T2T-CHM13 (Yang et al. 2023), 13 alleles (10.8%) resembled the T2T-CHM13 allele with a 15.4 kb deletion, while 45 alleles (37.5%) were similar to the GRCh38 allele without the deletion. Despite these discrepancies, we identified clinically important SVs consistently detected across both references. These include structural variation in immunoglobulin heavy chain (IGH) genes affecting antibody repertoire diversity (Rodriguez et al. 2023), mucin genes (*MUC1*, *MUC5AC*, *MUC7*, *MUC17*, *MUC21*, *MUC22*) involved in mucosal barrier function (Linden et al. 2008; Plender et al. 2024), and pharmacogenomic loci with possible implications for drug metabolism. Notably, a 5.8 kb heterozygous deletion in the *UGT1A* cluster affecting three genes (*UGT1A4*, *UGT1A5*, *UGT1A6*) was detected in 2 MENA individuals and was observed in only one South Asian individual across the entire 1K-GP dataset. Large deletions in UGT1A were highlighted previously to be responsible for Crigler-Najjar type I syndrome (Petit et al. 2008). A 2.2 kb heterozygous deletion in *CYP3A43*, detected in 3 MENA individuals (AF = 0.025), was predominantly carried by African populations (AFR AF = 0.266, AC = 143), with rare frequencies in the rest of the 1kGP continental populations. Additionally, we identified a 3.6 kb deletion in *CD55*, detected in 40 MENA samples and is a globally common polymorphism (global AF = 0.455), highlighting variation in this important gene, which is a target for malaria parasites and mediates their internalisation in erythrocytes (Shakya et al. 2021) (Figure S18). Additionally, the dual reference analysis identified a private germline heterozygous deletion affecting the TP53 coding sequence, a known cancer predisposition gene (Joerger et al. 2025) (Tables S24 and S25) (Supplementary note 4).

### MENA Structural variations intersection with OMIM exons reveal reference bias and population-specific alleles in disease-associated genes

We specifically investigated the intersection of MENA SVs with Online Mendelian Inheritance in Man (OMIM) exons (Amberger and Hamosh 2017). This overlap enabled the characterisation of population-specific SVs intersecting genes associated with OMIM phenotypes (n=4,866) (Hamosh et al. 2005). Analysis of GRCh38 SVs revealed 269 high-confidence unique SVs overlapping at least with one defined exon, affecting a total of 249 unique exons and 48 unique genes (Table S21). A parallel analysis was performed using T2T-CHM13 SVs lifted to GRCh38 coordinates, revealing 272 high-confidence SVs overlapping with the coding region of 53 genes. Genes demonstrating recurrent structural variations, *VPS53* and *LRPAP1*, showed similar patterns in both genomes, and, with 10 separate events each, though specific breakpoints varied (Tables S26 and S27).

Three SVs, a 457 bp tandem-duplication insertion in *ABCC1* (OMIM #618915), a 254 bp insertion in *XYLT1* (OMIM #615777) and a 110 bp insertion in *MED13L* (OMIM #616789), were nearly fixed and detected in 59 MENA samples. Concordantly, all three SVs are near-fication across all populations in the 1kGP ONT datasets. This observation suggests that these alleles are globally common and missing from the reference, being private to the GRCh38 genome (Table S26). In contrast, the T2T-CHM13-lifted SVs to the GRCh38 coordinates contain several high-frequency SVs overlapping with OMIM exons that were absent from the GRCh38 analysis. These SVs represent benign polymorphism and major alleles in MENA populations compared to the CHM13 reference, due to CHM13 reference bias, highlighting the need for reference-aware ancestry-specific variant interpretation guidelines in clinical genomics applications.

Furthermore, we observed a 3.8 kb deletion SV mapped to GRCh38, consistent with (– α 3.7 kb single gene deletion), the common α-thalassemia variant affecting 18 individuals (15 heterozygous and 3 homozygous). This deletion is common in MENA (Sniffles AF = 0.19) and enriched compared to the global population (Overall AF = 0.054), with the highest non-MENA frequency observed in AFR (AF 0.15), followed by AMR (0.035) and SAS (0.034), rare in EAS (0.008), while absent in EUR, consistent with the well-documented high prevalence of α-thalassemia in Middle Eastern, African, and Mediterranean populations (Figure S19) (Farashi and Harteveld 2018; Alhuthali et al. 2023). Long-read sequencing identified a pharmacogenomically critical 12.1 kb heterozygous deletion spanning the entire *CYP2*D6 (OMIM # 608902) exons in one individual, representing the *CYP2D6* null allele that eliminates drug metabolising capacity for approximately 25% of clinically prescribed medications. This deletion is globally rare (overall AF = 0.014), with the highest frequency in AFR (AF = 0.024 and AC = 13), indicating that *CYP2D6* whole-gene deletions are uncommon across all populations. Cytochrome P450 (CYP) genes are an important gene family for drug response and are challenging to sequence using short-read technologies, requiring specialised variant calling approaches (Lee et al. 2019).

Despite the healthy male cohort, multiple SVs were detected in X-linked genes, raising questions about pathogenicity classifications since most of these events are likely associated with a disease. SVs mapping to GRCh38 revealed nine distinct SHOX SVs with deletions ranging from 82-656 bp predominantly affecting exon 5, with the largest 656 bp VNTR deletion present in 36 individuals. This is a globally common VNTR polymorphism present across all populations (AF = 0.55). Given that SHOX haploinsufficiency is typically associated with short stature disorders, the global prevalence strongly suggests it may represent population-specific benign polymorphisms rather than pathogenic variants (Binder and Rappold 1993). Additionally, a large 19.7 kbp heterozygote deletion was detected in a female individual using both genome references intersecting with the 2^nd^ exon of *AMER1*. This SV is absent from the 1kGP ONT samples, representing a MENA private event. This is particularly important since *AMER1* frameshift mutations are associated with X-linked dominant condition Osteopathia Striata With Cranial Sclerosis, which usually affects females (Hague et al. 2017). The presence of these traditionally pathogenic X-linked SVs suggests either reduced penetrance or population-specific tolerance, highlighting the need for ancestry-specific interpretation of X-linked structural variants in clinical settings where MENA populations have been historically underrepresented in genomic databases.

### Combining MENA high confidence SVs with 1kGP SV catalogue and HGSVC2 SVs reduced SV burden in patients with MENA ancestries by up to 97.5%

To validate the clinical utility of the MENA high-confidence SVs catalogue, we assessed its effectiveness in reducing the burden of structural variations in 22 rare disease patients with negative whole-genome sequencing results, who were subsequently sequenced using Oxford Nanopore Technology. Across the 22 patients, a total of 675,703 SVs were identified, with an average of 30,713 SVs per individual, an average of 70 SVs per patient overlapping with OMIM exons, and an average of 4,797 SVs per patient overlapping with GIAB medically relevant genes. Due to the small sample size, filtering against the MENA high-confidence dataset alone using a 50% reciprocal overlap showed a minimal reduction in actionable SVs, reducing the total SV count by only 3.2%, with a similarly modest reduction in OMIM overlapping SV (0.2%) and medically relevant gene SVs (2.7%) (Table S28). However, when patients’ SVs were filtered against the dataset from 1,019 1k-ONT Sniffles calls merged with Sniffles called SVs from MENA individuals, a substantial reduction was observed, decreasing the total SV burden by 92%, with a corresponding reduction of 91% for OMIM-overlapping SVs (mean = 5.8), and 94% reduction for SVs overlapping medically relevant genes (mean = 281). The combined filtering approach using HGSVC2, 1K-ONT calls that includes MENA samples, achieved maximum reduction, decreasing the total SV burden in patients by 97.5% (Table S29).

### Long-read sequencing reveals population-level differences in clinically relevant repeat expansions between MENA and global populations

We compared Short Tandem Repeat (STR) lengths across 68 loci between the Mena dataset and the 1000 Genomes ONT Project dataset(Hiatt et al. 2024). For each individual, we examined both alleles and the shortest allele per locus. Our analysis focused on the 68 clinically relevant genes associated with human diseases and summarised in STRchive, previously assessed in the 1000 Genomes dataset (Hiatt et al. 2024; De Coster et al. 2024). Overall, STR length patterns were broadly similar between the two datasets (Figure 4C), though the MENA dataset showed lower amplitude, likely reflecting its smaller sample size compared to the 1000 Genomes project. Notably, STRs in the gene *EIF4A3* tended to have longer repeat units in MENA individuals compared with those from the 1000 Genomes dataset (Figure 4C, S17 and S23), although no alleles exceeded the pathogenicity threshold. In contrast, repeats in *ATXN3* were generally shorter in MENA individuals, and this difference became more pronounced when considering only the shortest allele per individual (Figure S20 and Table S28).

Regarding pathogenicity, 13 MENA individuals carried both alleles of *FOXL2* above the pathogenic threshold, compared to 50 in the 1kGP dataset. A single individual from the MENA cohort was found to have pathogenic repeat expansion in *PRNP*, when no individual in the 1000 Genomes dataset had alleles above the pathogenic threshold. However, due to the limited sample size of the MENA dataset, these differences should be interpreted with caution, as they are not representative of the broader population. Nevertheless, the STR repertoire of the MENA samples reported here adds to the catalogue of STRs present in healthy humans, which will allow for a more in-depth understanding of their effects on genes of medical interest.

### Pairwise SV-sharing between archaic humans and chimpanzee revealed ancient SVs in modern humans

To investigate SVs shared with archaic humans, archaic hominin genomes (three Neanderthals and one Denisovan) were genotyped against the modern human Sniffle-called SV callest using Paragraph (See method for details). Chimpanzee SVs were identified from previously sequenced chimpanzee using ONT (Soto et al. 2020), aligned against the GRCh38 and T2T-CHM13 genomes using Sniffles to identify SVs shared with modern humans.

Against the GRCh38, approximately 3 % SVs (5,517 SVs, affecting 1,833 protein-coding genes) were shared between humans and chimpanzees, indicating their origin predates the human-chimpanzee divergence approximately 6-7 million years ago (Glazko and Nei 2003). In addition, a total of 5.2% (9,722 SVs, affecting 2,467 protein-coding genes) are shared between modern humans and archaic hominins (Neanderthals and Denisovans), with Denisovan-shared SVs comprising the largest fraction at 3% (5,547 SVs, affecting 1675 protein-coding genes), followed by Altai Neanderthal variants at 2.3% (4,283 SVs, affecting 1,150 protein-coding genes) (Figure 5D and Table S30). Approximately 15.6% (28,946 SVs, affecting 4,666 protein-coding genes) appear to be modern human-specific, present across diverse global populations but absent from both chimpanzee and archaic hominin genomes, unique to Homo sapiens (Figure S19). Analysis using the CHM13 reference genome revealed a dramatic expansion in structural variant detection; however, the proportion of SVs shared with chimpanzee decreased to 1.5% against T2T-CHM13 (4,947 SVs, affecting 1,559 protein-coding genes). SV sharing pattern with archaic hominin reched 3.9% (13,014 SVs, affecting 2,188 protein-coding genes) (Table S30).

Analysis of SVs fixed across all populations (AF ≥ 0.95, MENA ≥ 0.90) identified 1,077 fixed variants in GRCh38 compared to 267 in T2T-CHM13, with the reduction reflecting removal of GRCh38 donor-specific reference bias. GRCh38 fixed variants showed remarkable evolutionary antiquity, with 46.2% (498 variants, affecting 237 protein-coding genes) shared with chimpanzee and 42.9% (462 variants, affecting 188 protein-coding genes) with archaic hominins. The CHM13 fixed variants showed comparable patterns, 36.0% chimpanzee-shared (96 variants) and 34.5% archaic-shared (92 variants), confirming these as deeply conserved structural elements maintained across primate evolution (Table S32).

## Discussion

This study introduces the first comprehensive catalogue of structural variants (SVs) in Middle Eastern and North African (MENA) populations, using published ultra-long Oxford Nanopore Technology (ONT) sequencing methods. By combining data from 61 individuals across eight MENA countries and applying a robust multi-caller approach against two human reference genomes (GRCh38 and T2T-CHM13), we explore the extensive population-specific genomic diversity and its importance for clinical genetics. Our dual-reference, multi-caller methodology demonstrates that comprehensive structural variant discovery can be achieved without the computational burden associated with de novo genome assembly or pangenome construction (Schloissnig et al. 2025; Beyter et al. 2021a; Gong et al. 2025a). Our dual-reference and multi-caller approach identified 97,765 high-confidence SVs with GRCh38 and 176,494 with T2T-CHM13, representing a twofold increase enabled by the complete telomere-to-telomere (T2T) assembly. The notable proportion of previously unknown variants (20.3% mapping to GRCh38) highlights the global underrepresentation of MENA genomes in current databases and emphasises the urgent need for ancestry-inclusive genomic resources.

This study demonstrates a significant advantage in structural variant (SV) discovery using the T2T-CHM13 reference genome. The results show a notable improvement in SV detection across the different algorithms, with T2T-CHM13 achieving a higher percentage of reads from total aligning to the genome, 95.2% compared to 88.4% for GRCh38, along with a greater number of detected variants (Chen et al. 2024). This improvement is especially evident in repetitive and segmentally duplicated regions, where traditional references like GRCh38 often show gaps or mis-assemblies. Importantly, the increased overlap of MENA SVs with repetitive sequences, 77.7% in T2T-CHM13 compared to 25.9% in GRCh38, highlights the ability of the gapless T2T-CHM13 genome to reveal previously inaccessible areas, including centromeric regions. The asymmetric success rates of coordinate lifting, ranging from 84% to 88% when moving from GRCh38 to CHM13, and only 34% to 43% in the reverse direction, further underline the enhanced mappability and representation of complex genomic regions in the CHM13 assembly (Chen et al. 2024; Paulin et al. 2025; Aganezov et al. 2022). In addition, this research extends these insights to populations that have traditionally been underrepresented, emphasising that reliance on incomplete reference genomes unfairly limits variant discovery in ancestries different from those of predominantly European donors. Therefore, the choice of a reference genome is not merely a technical decision but also a matter of equity in genomics, affecting which population-specific variants are detected in scientific studies and which remain hidden.

The availability of the 1kGP samples sequenced using ONT technology was important for contextualising MENA structural variants within global population diversity (Schloissnig et al. 2025). This ONT-based global reference enabled direct comparison using identical sequencing technology, eliminating platform-specific biases that would arise when comparing long-read MENA data to short-read global datasets.

Long-read sequencing technology enables the identification of complex structural variants (SVs) that are often missed by short-read methods. Our functional annotation analysis has revealed numerous gene-disrupting deletions, insertions, and duplications, many of which occur within genes of significant pharmacogenetic or clinical relevance. Notably, we identified a 12.1.1kb deletion covering all exons of *CYP2D6*, a null allele linked to poor metaboliser phenotypes across various drug classes. The structural variation found in genes such as *CYP3A43* and *UGT1A4-6* further reveals the unique pharmacogenomic landscape among MENA individuals, which may significantly impact drug effectiveness and toxicity. Additionally, deletions within the *CD55* gene, which interacts with malaria parasites, were present in over 60% of alleles, indicating a possible evolutionary adaptation to endemic infectious pressures (Shakya et al. 2021). Using our GRCh38 SVs set, we identified high-frequency variants intersecting OMIM genes that reveal systematic reference bias in clinical interpretation.

Long-read sequencing data for MENA individuals enabled the evaluation of repeat expansion in complex alleles difficult to assess using short-read technology. We identified repeat expansion amongst the 68 clinically relevant genes, associated with human diseases and summarised in STRchive, in MENA and 1K-GB samples that are difficult to fully interpret since individuals recruited for sequencing from MENA and in the 1K-GB are presumably healthy. These individuals may be carrying benign alleles of non-pathogenic expansion or may be at risk of developing disease later in life. Comparing MENA STR with the 1kGP dataset revealed population-level differences in repeat length distributions, particularly at the *EIF4A3* locus, where lengths were greater in individuals from the Middle East and North Africa (MENA) region, and shorter at the *ATXN3* locus. The pathogenic expansion in the 1KGB samples was speculated to be an artefact introduced during the cell culture process; however, since MENA samples were sequenced directly from donated blood, we rule out that presumption (Gustafson et al. 2024). Although the limited sample size warrants caution against overinterpretation, these findings imply that MENA populations might have distinct STR length spectra influenced by founder effects and demographic history. Since repeat expansions are involved in neurodevelopmental and neurodegenerative disorders, these results highlight the importance of extensive population-scale STR profiling. Such profiling could lead to the redefinition of pathogenic thresholds based on ancestry-specific distributions, rather than relying solely on universal cutoffs.

The integration of structural variants (SVs) from Middle Eastern and North African (MENA) populations with those from global and archaic datasets has revealed profound evolutionary patterns. Around 0.5 - 2.3-18% of MENA SVs show shared ancestry with Neanderthal and Denisovan genomes, while 1-2.3% also share ancestry with chimpanzee genomes, indicating their origins trace back to a period before the divergence of modern humans. The large number of archaic-shared SVs identified in the T2T-CHM13 reference genome highlights its improved ability to resolve ancient sequences absent in the GRCh38 reference. (Almarri et al. 2021). Furthermore, the notable decrease in the number of fixed variants from the GRCh38 reference (1,077) to the CHM13 reference (267) underscores that many “universal” SVs in GRCh38 are probably artefacts caused by reference bias, arising from private variants of the original reference donors. Consequently, analyses using complete and unbiased references like T2T-CHM13 offer a more precise view of human structural diversity, showing that truly fixed SVs are relatively few and highly ancient in evolutionary terms (Aganezov et al. 2022). Despite the small sample size and the UAE-biased, integration of MENA SVs with the 1k genomes ONT dataset proved essential for effective clinical variant filtering, achieving a 92.7% reduction in total SV burden, demonstrating that population-specific datasets are effective when integrated into broader genomic frameworks rather than used in isolation. This integration allowed us to accurately distinguish between common benign variants and truly rare potentially pathogenic variants in MENA patients. The maximum reduction was achieved by combining HGSVC2 with the global merged dataset and MENA SVs, reducing the total variant burden by 97.5% and filtering 93.5% of variants overlapping both OMIM exons and 95.3% of SVs in medically relevant genes, highlighting the synergistic value of integrating multiple population databases for comprehensive clinical variant interpretation.

The findings have important implications for equitable precision medicine implementation. For MENA populations marked by high consanguinity, complex admixture, and unique environmental pressures, the lack of representative reference data creates diagnostic blind spots where population-specific variants may be misclassified as pathogenic. Our high-confidence structural variant catalogue provides essential resources for improved variant annotation and reduced diagnostic inequalities. Incorporating MENA-specific variants into pharmacogenomic databases could enable regionally tailored drug dosing guidelines, while integration into pangenomic graphs and global resources will facilitate ancestry-aware variant interpretation. This study demonstrates that alignment-based approaches can efficiently generate comprehensive SV catalogues for underrepresented populations, revealing extensive population-specific genetic diversity while providing a cost-effective alternative to assembly-based methods. The substantial novel variation discovered, and its clinical implications, underscore the critical need for ancestry-inclusive genomic databases and interpretation guidelines, highlighting both the scientific necessity and ethical imperative of ensuring equitable representation of global genetic diversity in precision medicine applications.

## Methodology

### Quality Control and Read Assessment

Raw ONT data of 61 individuals from the Middle East and North Africa (MENA) were downloaded from the NCBI Sequence Read Archive (SRA). A total of 53 APR samples were sequenced using ultra-long read ONT sequencing as part of the UAE-based pangenome project available under the accession number PRJNA1108179 (Nassir et al. 2025). An additional eight KSA samples were sequenced as part of the Saudi and Japanese pangenome graph (JaSaPaGe) available under the accession PRJNA1091214 and PRJNA891101(Kulmanov et al. 2024, 2025). Sequencing of the obtained samples was performed using PromethION, and basecalling was performed using Guppy v5.1.13 with the super-accurate (SUP) model and Guppy (v6.5.7) with the High accuracy (HAC) model. Prior to Genome alignment and structural variant calling, the quality of Oxford Nanopore Technology (ONT) long-read sequencing data was comprehensively assessed using NanoPlot (v1.44.1) (De Coster and Rademakers 2023; Kulmanov et al. 2025). Quality metrics, including read length distributions, read quality scores, and N50 statistics, were generated for each sample to ensure data suitability for downstream structural variant analysis.

### Reference genome alignments

Reads were aligned to the GRCh38 and T2T-CHM13 linear reference genomes. For both reference genomes, we used minimap2 (v2.1.1) (Li 2018) to map the ONT reads using options tailored for ONT reads alignment “-a -x map-ont -L”. The alignment workflow involved creating reference genome indices using ‘minimap2 -d {reference} {reference.fasta}’, followed by alignment of ONT FASTQ files using “minimap2 -t 64 -a -x map-ont -L {reference}”. SAMtools (v1.21) was used to convert SAM to BAM format, sort the alignments by coordinates, and index the BAM file (Li et al. 2009; Danecek et al. 2021). Quality assessment metrics for the aligned BAM were generated using samtools flagstat for overall alignment statistics, samtools idxstats for per-chromosome read distribution analysis and coverage statistics were evaluated using cramino (v0.16.0) for both reference alignments “cramino {input.bam} --reference {reference.fasta}” (De Coster and Rademakers 2023).

### Multi-Algorithm SV Detection

Structural variants were identified using four complementary long-read SV calling algorithms to maximise sensitivity and accuracy. For each sample and reference genome combination, we employed Sniffles (v2.6.0) (Smolka et al. 2024; Sedlazeck et al. 2018), SVIM (v2.0.0) (Heller and Vingron 2019), Delly (v1.3.3) (Rausch et al. 2012), and CuteSV (v2.1.1) (Jiang et al. 2020). SVIM was executed in alignment mode with “--tandem_duplications_as_insertions and --interspersed_duplications_as_insertions” flags to capture complex rearrangements as insertions for improved downstream analysis. CuteSV was run with optimised clustering parameters: “--max_cluster_bias_INS 100”, “--diff_ratio_merging_INS 0.3”, “--max_cluster_bias_DEL 100”, and “--diff_ratio_merging_DEL 0.3”, along with “--genotype” flag for population-scale analysis. For SVIM and CuteSV, population-level merging was performed using JASMINE (v1.1.5) with “--normalize_type, --output_genotypes, --ignore_strand, --dup_to_ins”, and “--centroid_merging” parameters (Kirsche et al. 2023).

We used Sniffles to calculate candidate SVs and associated SNF files for each sample using standard parameters with reference genome support. We then employed Sniffles population-calling mode on all SNF files to generate multi-sample VCF files for both GRCh38 and T2T-CHM13 reference genomes (Smolka et al. 2024). Similarly, we used Delly’s population-calling approach optimised for long reads. We first called SVs per sample using the long-read (lr) subcommand, then merged all candidate SV sites using delly merge with options ‘-p -a 0.05 -v 3 -c’ to select PASS sites that are precise at single-nucleotide resolution with a minimum variant allele frequency of 5% and minimum coverage of 3×. We then genotyped this SV site list across all samples using delly lr and merged the results by ID using bcftools merge with -m id. We applied SANSA (v0.2.3) markdup subcommand with parameters ‘-y 0 -b 500 -s 0.5 -d 0.3 -c 0.1’ to remove redundant SV sites based on SV size ratio >0.5, maximum SV allele divergence of 30%, maximum breakpoint offset of 500 bp, and minimum fraction of shared SV carriers of 10% (Rausch et al. 2012; Danecek et al. 2021).

### Generating consensus calls supported by multiple callers

For each sample, structural variants called by the four algorithms were merged using JASMINE (v1.1.5) to create consensus call sets. The merging employed ‘--normalize_type --ignore_strand --dup_to_ins --centroid_merging –allow_intrasample’ parameters. This approach generates high-confidence structural variants supported by multiple independent calling algorithms. Consensus VCF files were filtered using BCFtools (v1.21) with ‘INFO/SUPP>=2’, ‘INFO/SUPP>=3’, and ‘INFO/SUPP>=4’ criteria to retain variants supported by multiple callers. Filtered single-sample consensus VCF files were merged across all samples using JASMINE (v1.1.5) with population-scale parameters including ‘--normalize_type --output_genotypes --ignore_strand --dup_to_ins. --centroid_merging’ The resulting multi-sample VCF files were processed using a custom script for VCF simplification to retain essential variant information, then sorted and indexed using bcftools (v1.21).

### Cross-reference coordinate lifting and consensus calls shared between GRCh38 and T2T-CHM13

To ensure high specificity and identify structural variants consistently detectable across reference genome assemblies, we generated consensus callsets of SVs shared between GRCh38 and CHM13 reference genomes for each algorithm and filtering threshold. We lifted the GRCh38 callsets to CHM13 coordinates using custom Python scripts to convert VCF files to BED format, followed by coordinate translation using UCSC liftOver tools with the GRCh38 to CHM13 chain file. We then compared the lifted VCF files with the original CHM13 VCF files to identify shared SVs using SANSA’s (v0.2.3) compvcf subcommand with options ‘-m 0 -b 50 -s 0.8’ to identify SVs with a size ratio ≥ 0.8 and a maximum breakpoint offset of 50 bp. For Delly callsets, we additionally applied the SV allele divergence filter -d 0.1 to require maximum SV allele divergence of 10%, leveraging Delly’s consensus SV allele sequences. As other algorithms lack local assembly information, the allele divergence filter was not applied to their callsets. Custom scripts then extracted true positive variants from the base VCF files to generate final consensus callsets representing structural variants consistently called across both reference genome assemblies. This cross-reference validation approach enhances callset reliability by identifying variants that are robust to reference genome choice and assembly differences. Additionally, the three-caller consensus SVs were specifically lifted using the “rtracklayer” R package, achieving a slightly better lifting efficiency compared to UCSC’s liftOver tool for both GRCh38 and T2T-CHM13 SVs. 91.8 % of GRCh38 SVs were lifted using this method to T2T-CHM13, and 49.2 % were lifted reciprocally.

### Concordance of SV calling between different long-read callers and discovery of Novel SVs

To identify concordance between different long-read SV calling algorithms employed in this study and between each long-read caller and three-caller consensus callsets, we employed Truvari (v5.3.0) bench for comprehensive evaluation of SV calling concordance. We conducted pairwise comparisons between all four calling algorithms (n=6 combinations) and systematic evaluation of each caller against consensus callsets using ‘--passonly’ filtering.

To identify novel structural variants within our MENA cohort, we systematically compared our nanopore-derived three-caller consensus SV call set against established public repositories. We applied a minimum threshold of 50% reciprocal overlap to determine whether SVs in our dataset had been previously documented in publicly available structural variant databases. MENA SVs were compared to SV databases from diverse populations, including gnomAD-SV(Collins et al. 2020) (v2.1), representing high-coverage short-read sequencing data from 14,891 globally diverse samples; the Audano et al.(Audano et al. 2019) dataset, derived from long-read sequencing of population-diverse samples; the Human Genome Structural Variation Consortium (HGSVC2) (Ebert et al. 2021) collection; and dbVar (Lappalainen et al. 2013), which aggregates structural variants from multiple studies across different sequencing platforms. Additionally, we compared MENA SVs to population-scale ONT sequencing specific populations, including the SVs dataset generated from sequencing 945 Han (Gong et al. 2025a) individuals and the SV catalogue generated from ONT sequencing of 3,622 Icelanders(Beyter et al. 2021a). For the GRCh38 reference SVs and the T2T-CHM13 SVs lifted to GRCh38 reference coordinates, SV overlap assessment was performed using bedtools intersect (v2.31.1) with the ‘-f 0.5 -r’ parameters to enforce bidirectional 50% reciprocal overlap. Structural variants meeting this threshold were classified as previously reported, while those falling below this criterion were designated as novel discoveries.

### Structural variants filtering and annotation

Following consensus callset generation, we applied stringent quality filtering to retain only PASS variants with size ≥ 50 bp using BCFtools (v1.21) filter “(INFO/SVLEN >= 50 || INFO/SVLEN <= -50) || (INFO/END - POS >= 50)” and restricted analysis to standard chromosomes (chr1-22, chrX, chrY) to remove technical artefacts from unplaced scaffolds and alternative haplotypes. We calculated comprehensive population genetic statistics using BCFtools +fill-tags plugin, generating INFO fields including allele count (AC), allele frequency (AF), total alleles (AN), sample count (NS), missing genotype fraction (F_MISSING), heterozygous/homozygous counts (AC_Het, AC_Hom), Hardy-Weinberg equilibrium (HWE), excess heterozygosity (ExcHet), and minor allele frequency (MAF). These statistics enabled systematic characterisation of structural variant distribution patterns, quantification by variant type and sample, and size distribution analysis across the MENA population cohort. Finally, we performed functional annotation using Variant Effect Predictor (VEP)(McLaren et al. 2016) “--offline” with regulatory feature annotation “--regulatory” for both GRCh38 and T2T-CHM13 assemblies. Functional impacts, clinically relevant and protein-affecting structural variants were identified against both reference genomes. Protein-affecting variants is a broader term we adopted, encompassing frameshift variants, in-frame deletions and insertions, stop-gained/lost mutations, start-gained variants, protein-altering variants, transcript deletion and amplification events, and coding sequence variants that collectively represent structural changes with a direct impact on protein sequence. Additionally, SVs were further annotated using Structural Variants Annotator (SVAN) using default parameters to classify SVs into distinct classes, including Mobile Element Insertions (MEI), processed pseudogene integrations, various forms of duplications, tandem repeats expansions/contractions and nuclear-mitochondrial segments (NUMT) (Schloissnig et al. 2025).

### SVs’ intersections with gene regions, GiaB startifications, repeat regions according to RepeatMakser annotation and OMIM exons

MENA SVs intersecting Genome in a Bottle (GiaB) stratification, gene regions and repeat regions were identified using overlaps (Dwarshuis et al. 2024). Any function provided by the GenomicRanges R package. The most updated GiaB stratifications (v3.6) were downloaded from the GiaB FTP server https://github.com/usnistgov/giab-stratifications?tab=readme-ov-file. Intersection of MENA SVs with several stratifications was investigated, including segmental duplication using the “segdups.bed.gz” file, all difficult regions using the “alldifficultregions.bed”, high-confidence regions using the “notinalldifficultregions.bed”, and refseq coding region using the “refseq_cds.bed.gz. To investigate GRCh38 SVs overlapping with gene regions, we used gene regions from the geocode annotation (Harrow et al. 2012), while for the T2T-CHM13 SVs, the intersection was done using the gene regions from “chm13.draft_v2.0.gene_annotation.gff3” (Nurk et al. 2022). To investigate SVs intersecting repeat regions, RepeatMasker track for GRCH38 was downloaded from the UCSC Table Browser and RepeatMasker bed file was downloaded from the T2T-CHM13 GitHub page https://github.com/marbl/CHM13. SVs inserting centromeric regions in GRCh38 were identified using the centromere track downloaded from the UCSC table browser, while centromeric regions for T2T-CHM13 were downloaded from the T2T-CHM13 GitHub page https://github.com/marbl/CHM13. Intersection between OMIM exons and MENA GRCh38 SVs was identified using bedtools intersect to find any overlap between MENA SVs and OMIM exons using the “-wa -wb” option. CHM13 SVs lifted to GRCh38 were used to investigate the intersection of CHM13 SVs with OMIM exons.

### Reduction of actionable structural variants in the rare disease patient cohort

Structural variants files from a set of undiagnosed patients (N = 22) of mostly Arab ancestry, who previously had unresolved testing using short-read whole-exome sequencing (WES) and were subsequently sequenced using ONT (Sinha et al. 2025), were downloaded from https://figshare.com/s/e58cf877382d23b6b6db. To filter patient SVs using different SV catalogues, we built a user–friendly web–browser–based application available at github link: https://github.com/BackofenLab/SV_diagnostic_tool. In summary, the tool in the background performs normalisation and indexing using bcftools (v1.19), followed by variant comparison with Truvari (v5.4.0) using user-defined thresholds. In our analysis, we used a threshold of 0.5 reciprocal overlap to determine if patients’ SVs had been previously reported in a defined database. The workflow compares patients’ structural variants with SV catalogues from HGVSC2, 1k-ONT project sniffles calls merged with MENA SVs called using Sniffles, and three-caller-consensus MENA SVs. The tool evaluates overlap with medically relevant genes and OMIM exons and intersects results with a global database of known structural variants. All analyses are presented with visualisations generated with the matplotlib library.

### STR expansion genotyping

Short tandem repeats (STRs) are highly mutable repetitive sequences associated with over 40 neurological and developmental disorders (Fan and Chu 2007). Long-read sequencing enables accurate STR characterisation by spanning entire repeat regions, overcoming PCR biases and size limitations inherent to short-read approaches. We employed STRdust, a tool designed to address the high error rate of long-read sequencing while detecting multiple STRs within single reads, for targeted genotyping of pathogenic STR loci. STRdust analysis was performed on GRCh38-aligned BAM files with the following command “STRdust --pathogenic --threads 16 --sample {sample_name} --sorted –unphased {reference.fasta} {input.bam} | bgzip > {output.vcf.gz}”. This approach enabled systematic screening for pathogenic STR expansions across the MENA cohort, complementing structural variant analysis with targeted assessment of repeat expansion disorders. We downloaded the dataset associated with the 1000 Genomes Project from the pathSTR website (https://pathstr.bioinf.be/), which integrates data from Noyvert et al. (2023), Schloissnig et al. (2024), and Gustafson et al. (2024). The dataset was filtered to retain only entries corresponding to “STRdust_hg38,” ensuring comparability with our STR dataset. Our STR loci were assigned to genes when located within 50 bp of the annotated start or end of a gene. For consistency, we focused our analysis on the 68 clinically relevant genes previously analysed in the 1000 Genomes dataset and compared the distribution of allele lengths (in repeat units) between datasets. To assess potential pathogenicity, we obtained the list of disease-associated STR loci from STRchive (“https://raw.githubusercontent.com/dashnowlab/STRchive/refs/heads/main/data/catalogs/ST Rchive-disease-loci.hg38.general.bed”). This resource provided, for each gene, the minimum number of repeat units associated with pathogenicity, as well as the reference motif length (i.e., the size of the repeated sequence unit).

### Merging MENA SVs with the 1k ONT SVs set

To contextualise MENA structural variants within a broader population framework, we merged our Sniffles callsets with publicly available structural variants from the 1000 Genomes Project ONT data called using Sniffles for both GRCh38 and T2T-CHM13. Due to computational constraints with large multi-sample VCF files, we employed a chromosome-stratified merging approach. Individual sample VCF files from both studies were split by chromosome, creating separate VCF files for each of the 24 standard chromosomes plus mitochondrial DNA. SURVIVOR (v1.0.7) was then applied independently to each chromosome with parameters optimised for long-read data: 500 bp maximum distance between breakpoints, minimum of 1 supporting caller, consideration of variant type and strand information, and minimum variant size of 50 bp ‘SURVIVOR merge {file_list.txt} 500 1 1 1 1 50 {chr_merged.vcf}’. Following per-chromosome merging, we merged the per-chromosome VCF by chromosomes to produce the final 1K SVs plus MENA VCF. We applied custom Python scripts to simplify the merged VCF files by retaining only essential fields (SUPP, SVTYPE, SVMETHOD, END, STRANDS, AVGLEN, CIPOS, CIEND) and FORMAT fields (GT, LN, DR) while removing memory-intensive fields (SUPP_VEC, PSV) to enable efficient downstream analysis. Despite merging SVs called by the same caller (Sniffles) with the 1kGP ONT sequencing, 39% (33,958 SVs) of GRCh38 SVs and 50% (104,880) of T2T-CHM13 SVs didn’t merge with the 1K samples, highlighting the ongoing challenge in the field when merging SVs across different studies due to breakpoint differences.

PCA was run separately for the GRCh38 and CHM13 SV datasets using PLINK2, requesting 10 principal components per reference. Input SV sets were generated by applying length filtering (50 bp ≤ SV length < 10,000 bp; using AVGLEN for insertions and abs(END − POS) for deletions, duplications and inversions) and retaining samples present in both the VCF and sample_population_table.txt. Before PLINK import, VCFs were filtered to retain variants marked as passing upstream filters in the VCF FILTER field, remove sites with no called genotype, and retain polymorphic variants with minor allele count ≥1 (bcftools view -f PASS -U -c 1:minor). During PLINK import, variants were restricted to autosomes (--autosome), VCF half-calls were treated as missing (--vcf-half-call missing), and variants missing in more than 5% of samples were excluded (--geno 0.05). VCFs were converted into PLINK2 genotype files (--double-id, using each VCF sample name as both family and individual ID; --make-pgen, producing .pgen/.pvar/.psam files), and PCA was run with --pca 10.

Differentiated SVs (Fst ≥ 0.2) in MENA were identified using VCFtools per-site and window-size (1 Mb window) *Fst* between MENA and the rest of the samples from the 1K ONT. To assess allele frequency concordance between MENA and global populations, Pearson correlation coefficients were calculated between MENA and each continental population (AFR, AMR, EAS, EUR, SAS) across all shared SV sites in the merged dataset. SVs were filtered to retain variants with AF ≥ 0.01 and AF ≤ 0.95 in each compared population, and AF ≥ 0.01 and AF ≤ 0.90 in MENA, excluding rare variants and near-fixed alleles likely reflecting reference bias.

### Archaic Hominin SV Genotyping and Chimpanzee SV Calling

To investigate structural variant sharing between modern MENA populations and archaic hominins, we genotyped four archaic hominin genomes (Altai Neanderthal(Prüfer et al. 2014), Chagyrskaya Neanderthal (Mafessoni et al. 2020), Vindija Neanderthal (Prüfer et al. 2017), and Denisovan (Petr et al. 2020) using population-scale SV callsets. BAM files were obtained from the Max Planck Institute for Evolutionary Anthropology (https://www.eva.mpg.de/genetics/genome-projects/) and converted to FASTQ format using SAMtools (v1.21). Due to the degraded nature of ancient DNA, alignments were performed using BWA-MEM (v0.7.17) with parameters optimised for ancient DNA: reduced seed length (-k 12), reduced minimum seed length (-W 20), reduced re-seeding rate (-r 1.0), and adjusted mismatch and gap penalties (-A 1 -B 1) to accommodate ancient DNA damage patterns. Alignments were generated against both GRCh38 and T2T-CHM13 reference genomes, followed by duplicate marking using GATK (v4.6.1.0) MarkDuplicates with --REMOVE_DUPLICATES false to retain all informative reads while flagging potential PCR duplicates. Alignment rate for archaic samples is available in Table S31.

SV genotyping was performed using Paragraph (v2.3) using default parameters with population-scale VCF files as input, containing structural variants identified from the merged MENA and 1000 Genomes Project cohorts. Genotyping was conducted independently for GRCh38 and T2T-CHM13 aligned samples to enable cross-reference validation of archaic hominin genotypes. For comparative evolutionary analysis, we additionally called structural variants from a chimpanzee sample sequenced using Oxford Nanopore Technology (Soto et al. 2020). Following the same analytical pipeline employed for MENA samples, chimpanzee reads were aligned to both GRCh38 and T2T-CHM13 human reference genomes using minimap2 (v2.1.1) with ONT-specific parameters, and structural variants were called using Sniffles (v2.6.0). This approach enabled the identification of structural variants shared between humans and chimpanzees, providing evolutionary context for modern human structural variation.

## Data Availability

All data produced in the present work are contained in the manuscript

https://www.ncbi.nlm.nih.gov/bioproject/PRJNA1108179

https://www.ncbi.nlm.nih.gov/bioproject/PRJNA1091214

https://www.ncbi.nlm.nih.gov/bioproject/PRJNA891101

https://figshare.com/s/e58cf877382d23b6b6db

https://www.eva.mpg.de/genetics/genome-projects/

## Author Contribution Statement

T.A.Y. contributed to conceptualisation, data curation, formal analysis, investigation, methodology, project administration, resources, validation, visualisation, writing, original draft preparation, and writing and editing the manuscript. S.T. contributed to the investigation, methodology, bioinformatics pipelines, visualisation, and writing and editing the manuscript. S.H. contributed to the investigation, methodology, and reviewed the manuscript. S.A. contributed to reviewing and editing the manuscript. P.H. contributed to reviewing and editing the manuscript. R.B. contributed to reviewing and editing the manuscript. A.A.T. contributed to the investigation, reviewing and editing the manuscript. O.S.A. contributed to conceptualisation, funding acquisition, investigation, and writing and editing the manuscript.

## Code and data availability

VCF file generated in this study are available in https://doi.org/10.5281/zenodo.18620536. Scripts and R-scripts used in this study are available at https://github.com/alyazeeditalal/MENA-SV-discovery. While the SV diagnostic tool designed to reduce actionable SVs in this study is available at https://github.com/BackofenLab/SV_diagnostic_tool.

## Acknowledgment

We would like to acknowledge that the discussion we had with Marc Haber (Birmingham University, Birmingham, UK) and Mohammed Almarri (Dubai Police, Dubai, UAE) has significantly helped to shape this research study. The author thanks MBRU IT, which maintained the computer cluster on which the data were hosted, and the analyses in this study were performed.

