## Supplemental tables for "Structural variation landscape of Middle Eastern and North African individuals from long-read nanopore sequencing reveals medically relevant variants"

### Supplementary note 1

#### Identifying SVs using multi-callers against GRCh38 and T2T-CHM13 reference genomes

We conducted a comprehensive SV discovery against GRCh38 and T2T-CHM13 reference genomes using four long-read-based SV callers: CuteSV (Jiang et al. 2020), Delly (Rausch et al. 2012), Sniffles (Smolka et al. 2024), and SVIM (Heller and Vingron 2019). Individual callers identified between 94,976-126,464 SV sites for GRCh38 and between 183,834-268,409 SV sites for T2T-CHM13, with T2T-CHM13 consistently enabling higher detection than GRCh38 across all tools (Figure S2 and S3) (Table S4). To reduce false positive calls, we selected the three-caller consensus merge approach, where an SV should be called by at least three callers, while avoiding the deletion-elimination bias observed when we merged by four-caller consensus (SV called by all four callers) (Details in Method) (Table S5). The 4-caller consensus eliminated virtually all deletions, indicating fundamental algorithmic differences in deletion calling between callers that prevent unanimous agreement despite high insertion concordance. Consensus calling requiring an SV to be called by at least three callers yielded, in total, 97,765 (GRCh38) and 176,494 (T2T-CHM13) high-confidence SVs. GRCh38 three-caller consensus SVs are divided into 42,402 deletions (43.4%), 55,301 insertions (56.6%) and 62 duplications (0.06%). On the other hand, the T2T-CHM13 three-caller consensus SVs encompass 79,259 insertions (44.9%), 97,185 deletions (55.1%) and 50 duplications (0.03%) (Figure S4).

Pairwise concordance analysis using Truvari bench (English et al. 2022) revealed varying caller complementarity. CuteSV vs SVIM showed the highest overlap for GRCh38 shared called SVs, constituting 80.8% and 79.7% of total calls, respectively. In contrast, Sniffles vs SVIM performed best for T2T-CHM13, where shared SVs constituted 69.8% and 67.7% of total calls for each tool, respectively. Delly consistently showed lower concordance with other callers across both references (Figure 1 I and J) (Tables S9). When we investigated callers' concordance with the high-confidence three-caller call set, SVIM showed the highest percentage overlap, 95.1% of the total reads for both consensus SVs against the GRCh38 and the T2T-CHM13 (Tables S10). Reciprocal coordinate lifting between GRCh38 and T2T-CHM13 call sets using stringent overlap criteria (size ratio ≥ 0.8, maximum breakpoint offset of 50 bp) revealed asymmetric lifting efficiency with a higher percentage of SVs lifted from GRCh38 (84.2-88.4% across callers) to T2T-CHM13 versus 34.1-42.7% in the reverse direction (Table S11). This asymmetric lifting efficiency reflects the improved representation of complex genomic regions in T2T-CHM13 that cannot be mapped back to the incomplete GRCh38 assembly(Paulin et al. 2025; Aganezov et al. 2022). After reciprocal validation, we identified a consensus set of SVs for each tool that were called against both genomes in both GRCh38 and T2T-CHM13 coordinates (Table S12).

To identify robust SVs that can be detected across different reference genomes, we performed reciprocal coordinate lifting between GRCh38 and T2T-CHM13 call sets. GRCh38 SV coordinates across all callers were lifted to T2T-CHM13 using UCSC's liftOver tool, and the lifted coordinates were compared against the original T2T-CHM13 calls to identify shared SVs. Similarly, T2T-CHM13 coordinates were lifted to GRCh38 for reciprocal validation. Shared SVs called against both genomes were identified using stringent overlap criteria requiring a size ratio ≥ 0.8, a maximum breakpoint offset of 50 bp to account for the inherent differences in SV representation between reference genomes (See methods for details). Cross-reference coordinate lifting revealed substantial genome-specific SV detection, with liftOver efficiency from GRCh38 to T2T-CHM13 ranging from 84.2-88.4% across callers, while lifting from T2T-CHM13 to GRCh38 showed lower success rates (34.1-42.7%). This asymmetric lifting efficiency reflects the improved representation of complex genomic regions in T2T-CHM13 that cannot be mapped back to the incomplete GRCh38 assembly(Paulin et al. 2025; Aganezov et al. 2022). After reciprocal validation, we identified a consensus set of SVs for each tool that were called against both genomes in both GRCh38 and T2T-CHM13 coordinates (Table S11). The three-caller consensus achieved the highest cross-genome consistency, with 46.1% of total SVs identified against GRCh38 also identified in the T2T-CHM13 (Table S12). This approach identified the most robust MENA SV call set, consisting of SVs that can be reliably detected regardless of the reference genome choice, providing a high-confidence foundation for population-level structural variant analysis.

**Supplementary note 2**

**Allele frequency and size distribution of MENA SVs**

Allele count (AC) and allele frequency (AF) of SVs discovered against both reference genomes revealed distinct patterns. For GRCh38, 28.86% (28,129) of variants were singletons (AC = 1), 11.81% (11,510) were doubletons (AC = 2), and 59.33% (57,819) were polymorphic (AC ≥ 3), with 93.2% (91,155) classified as common (MAF > 5%) and 8.4% (8,248) nearly fixed (AF ≥ 90%). Due to the inclusion of the previously unresolved regions in T2T-CHM13 and the reduced reference bias exposing additional individual-specific variants, the T2T-CHM13 reference genome showed a higher proportion of singletons at 39.27% (69,127), and doubletons at 15.99% (28,154), with 44.74% (78,767) polymorphic variants and 7.8% (13,816) nearly fixed variants (AF ≥ 90%), though maintaining similar common variant frequency at 93% (164,066, MAF > 5%) (Table S13). T2T-CHM13 consistently detected larger SVs compared to GRCh38, with greater mean sizes for both deletions (988 bp vs 768 bp) and insertions (513 bp vs 474 bp), impacting larger genomic regions per sample (12.2 Mb vs 11.6 Mb) (Table S7 and S8 and Figure S6 and S7). Small SVs (<1 kb) predominated in both datasets (89.1% GRCh38; 82.5% T2T-CHM13), though T2T-CHM13 detected a higher proportion of large SVs (≥1 kb) at 17.5% compared to 10.9% in GRCh38 (Table S14).

**Supplemental note 3**

**Structural variation class distribution and characterisation**

We conducted a comprehensive analysis of structural variations in the MENA population using the Structural Variant Annotator (SVAN), an algorithm that annotates and classifies sequence-resolved insertions and deletions into distinct classes, leveraging their allelic representations and genomic annotations. SVAN successfully classified the majority of structural variations in our resources, offering a detailed perspective of SV classes across both genomes(Schloissnig et al. 2025).

SVAN classified 51,398 insertions (92.9% of total insertions) and 22,966 deletions (54.2% of total deletions) from our GRCh38 3-caller SVs dataset; in total, 74,364 SVs were annotated using SVAN (76.0% of all SVs) (Figure S8). A total of 9,245 (9.5%) of all SVs were classified as duplications, of which 7,461 (80.7%) represent tandem duplications, while 1,618 (17.5%) constitute interspersed duplications. We additionally identified 81 inverted duplications (0.9%) and 23 complex duplications (0.2%). Additionally, 50,863 SVs (52.0% of total SVs) are classified as variable number tandem repeats (VNTRs), with insertions (31,457) substantially exceeding deletions (19,406). Then we focused on the distinct classes of insertions classified by SVAN, most of which reflect mobile element activity. These include 12,141 non-reference mobile element insertions (MEIs), comprising 10,487 Alu, 1,632 L1, and 922 SVA insertions. We further identified 2,875 deletion events of reference MEIs. We also identify 41 non-reference processed pseudogenes and 26 reference polymorphic processed pseudogenes. We further observe evidence for endogenous retroviral activity, with 31 insertions classified as HERVK and 25 solo-long terminal repeats (LTRs). Finally, SVAN classifies 34 insertions as nuclear mitochondrial DNA segments (NUMTs) with a median length of 640 bp (Figure S8).

Classification of SVs identified using the CHM13 reference with SVAN successfully classified 73,129 insertions (92.3% of total insertions) and 63,640 deletions (65.5% of total deletions), representing 136,769 classified variants (77.5% of all SVs). The CHM13 analysis revealed 14,218 duplications (8.1% of total SVs): 11,318 tandem duplications (79.6%), 2,741 interspersed duplications (19.3%), 86 inverted duplications (0.6%), and 23 complex duplications (0.2%). A subset of 50 (0.4%) duplication events was identified using the long-read caller algorithms implemented in our SV discovery pipeline. VNTRs dominated the CHM13 classification with 112,004 variants (63.5% of total SVs).  None-reference mobile element insertions in CHM13 revealed 10,151 total MEIs, including 8,023 Alu, 1,239 L1, and 660 SVA insertions. We identified 39 NUMTs with a median length of 1,026 bp and 36 processed pseudogenes, indicating robust detection of these rare insertion classes. The comprehensive SVAN classification across both reference assemblies provides unprecedented resolution of SV architecture in MENA populations and highlights the critical importance of the reference genome in SV classes classification (Figure S9).

**Supplementary note 4**

**Reference genome bias in discrepant regions.**

The reference genome bias was significantly evident when inspecting the functional consequences of SVs at discrepant regions GRCh38 and T2T-CHM13 references. Genes in discrepant regions between GRCh38 and T2T-CHM13 revealed a different pattern of structural variations in MENA individuals based on the underlying reference used. When GRCh38 was used as a reference, a whole transcript deletion (1.84 kbp deletion) in *GSTM1* affecting 6 transcripts of this gene was detected in 25 individuals (21 heterozygous, 4 homozygous deletions) (Figure S10). *GSTM1* is a glutathione S-transferase detoxification enzyme belonging to the mu family, important for kidney disease progression and involved in the detoxification of electrophilic compounds, including carcinogens, therapeutic drugs, and environmental toxins (Gigliotti et al. 2020). Contextualising this variant against the 1K-ONT dataset, the *GSTM1* deletion is common in MENA (Sniffles AF = 0.40) and enriched compared to the global frequency (overall AF = 0.07), with the highest non-MENA frequency observed in EUR (0.14), followed by AMR (0.09), SAS (0.06), EAS (0.04), and AFR (0.03). The elevated frequency in MENA and European populations is consistent with published literature reporting high *GSTM1*-null prevalence in these ancestries. *GSTM1* is depleted in the T2T-CHM13 by a ~17 kbp deletion in comparison with the GRCh38 reference (Saitou et al. 2018; Yang et al. 2023); hence, we detected a 1.84 kbp insertion in at least one MENA sample when aligned to the T2T-CHM13. The deletion we observed against the GRCh38 indicates that these samples resemble the T2T-CHM13 deleted allele of *GSTM1*. We also detected a heterozygous ~18.5 kbp deletion intersecting with *CR1* exons using the GRCh38 as a reference, which was not detected when the T2T-CHM13 was used as a reference (Figure S11). This deletion is common in MENA (Sniffles AF = 0.14) but rare globally (overall AF = 0.005), with low-frequency or rare occurrences across all non-MENA populations. This is a known discrepant region observed in T2T-CHM13, resulting in the depletion of eight exons (450 amino acids) in *CR1(Yang et al. 2023)*. Additionally, the KLRC region harbours a ~15.4 kbp structural discrepancy between GRCh38 and T2T-CHM13 reference genomes, where KLRC2 is deleted in T2T-CHM13 (Yang et al. 2023). When individuals were mapped to GRCh38, we detected 12 heterozygous 15.4 kbp deletions, where one copy resembles the T2T-CHM13 allele (Figure S12). This deletion is common in MENA (Sniffles AF = 0.12) and low-frequency globally (overall AF = 0.04). Conversely, when the same cohort was mapped to CHM13, a 15.4 Kbp insertion was detected, with 18 samples homozygous and 9 samples heterozygous for the additional sequence absent in CHM13. Our dual reference approach enabled the detection of the KLRC2 haplotypes in MENA individuals, where 45 alleles (37.5%) are GRCh38-like, containing the 15.4 kb sequence, while 13 alleles (10.8%) are CHM13-like, lacking this insertion, and the remaining 62 alleles (51.7%) could not be confidently genotyped. Using the T2T-CHM13 as a genome reference resolves the mapping error observed in the MAP2K3 gene (Chen et al. 2024). When the GRCh38 was used as a reference, a whole heterozygous transcript deletion of 8.2 kbp was detected in MAP2K3 affecting 51 individuals due to a mapping error, which was not detected when the T2T-CHM13 reference was used (Figure S13 and S14).

Despite the discrepancies between GRCh38 and T2T-CHM13 genomes, we identified functionally consequential SVs of clinical and medical importance matched in both reference genomes. We found systematic structural variations affecting immunoglobulin heavy chain (IGH) transcripts in both reference genomes, highlighting the diversity in MENA population immunogenetics. The discovered IGH polymorphism in the MENA population impacts naive and antigen-experienced antibody repertoire, indicating qualitative and quantitative differences in antibody response (Rodriguez et al. 2023). Detected transcripts disruptions in both genomes in IGH genes include *IGHV7-4-1* transcript deletion (9.6 kbp deletion detected in 34 samples), which is common in MENA (Sniffles AF = 0.42) and globally (overall AF = 0.35), with the highest frequency in AFR (0.54). An 11.8 Kbp deletion in one individual affecting *IGHV4-61*, *IGHVII-60-1* and *IGHV3-60* transcripts was rare in both MENA (Sniffles AF = 0.008) and globally (overall AF = 0.004). A frameshift disruption in IGHG1 caused by a 565bp insertion was detected in 1 sample, and three deletions in *IGHG3* (186-188bp deletions detected in 1-8 samples) (Table S19 and S20).

Furthermore, we detected significant structural diversity using both reference genomes in mucosal barrier genes that play critical defensive roles in pathogen entrapment and mucociliary clearance(Linden et al. 2008). This structural diversity includes a 129 bp coding sequence deletion in *MUC1* affecting two individuals, a 69 bp in-frame deletion in *MUC7* detected in 6 individuals, a 2.7kb insertion in *MUC17* detected in 14 individuals, 90 bp in-frame insertion in *MUC21* detected in 14 individuals, 1074 bp in-frame deletion in *MUC22* affecting 11 individuals, a 192bp insertion in *MUC5AC* detected in 42 samples (Plender et al. 2024). We observed critical structural variants in drug metabolism genes detected on both genome references, including a 2.2kb deletion affecting *CYP3A43* coding sequence, detected in 3 samples. This deletion is low-frequency in MENA (Sniffles AF = 0.025) but common globally (overall AF = 0.08), driven primarily by the high frequency in AFR (0.27), while rare to low-frequency in other populations (AMR = 0.04, SAS = 0.01, EUR = 0.006, EAS = 0.003). A 5.8kb deletion in the *UGT1A* cluster was detected in two individuals, affecting the coding sequence of 3 genes from this cluster (*UGT1A4*, *5* and *6*). This deletion is low-frequency in MENA (Sniffles AF = 0.017) and rare globally (overall AF = 0.0005), with the only non-MENA detection in SAS (AF = 0.003). Large deletions in *UGT1A* were highlighted previously to be responsible for Crigler-Najjar type I syndrome (Petit et al. 2008). Additionally, we identified a 3.6kb deletion detected in 40 samples in CD55 (25 heterozygous deletions and 15 were homozygote), highlighting the structural diversity of this important gene that is a target for malaria parasites and mediates their internalisation in erythrocytes(Shakya et al. 2021) (Figure S15 and S16). This deletion is common in MENA (Sniffles AF = 0.51) and globally (overall AF = 0.45) and may potentially confer protection against Malaria parasite infection. Additionally, the dual reference analysis allowed the detection of a private 558bp germline heterozygous deletion affecting the TP53 coding sequence, a known cancer predisposition risk gene (Joerger et al. 2025) (Table S19 and S20).

**Figure S1. Quality assessment and alignment statistics for MENA ONT sequencing data.** Violin plots showing (**A**) N50 read length values across 61 MENA samples with a mean of 56 kb, (**B**) total sequencing output per sample, (**C**) Distribution of the longest read sequenced per sample, and (**D**) Distribution of sequenced reads' mean quality for samples included in this study. (**E**) The read quality score distribution was plotted as a box plot; the number of reads for each category is shown on the y-axis. The total number of reads and the percentage of alignment from total reads aligning to GRCh38 are represented in violin plots in (**F**) and (**G**), respectively. Total reads and the percentage of reads that align to the T2T-CHM13 reference genome are shown in violin plots in (**H**) and (**I**), respectively. Mean coverage per sample for all samples included in our study is illustrated in barplots for GRCh38 (**J**) and T2T-CHM13 (**K**).

**Figure S2. SV detection performance by individual callers on the GRCh38 reference.** Bar plots showing the distribution of SV counts per sample for each caller: (**A**) CuteSV, (**B**) Delly, (**C**) Sniffles, and (**D**) SVIM. Each plot displays deletions (Green), insertions (Blue), inversions (Pink) and duplications (Orange). Size distribution of insertions (Blue) and deletions (Green) in two separate windows: ≥ 1kb and ≤ 1kb for each caller (**E**) CuteSV, (**F**) Sniffles, (**G**) SVIM, and (**H**) Delly. A peak at size 300bp and 6000bp is detected across all callers, representing Alu and LINE retrotransposons, respectively.

**Figure S3. SV detection performance by individual callers on the T2T-CHM13 reference.** Bar plots showing the distribution of SV counts per sample for each caller: (**A**) CuteSV, (**B**) Delly, (**C**) Sniffles, and (**D**) SVIM. Each plot displays deletions (Green), insertions (Blue), inversions (Pink) and duplications (Orange). Size distribution of insertions (Blue) and deletions (Green) in two separate windows: ≥ 1kb and ≤ 1kb for each caller (**E**) CuteSV, (**F**) Sniffles, (**G**) SVIM, and (**H**) Delly. A peak at size 300bp and 6000bp is detected across all callers, representing Alu and LINE retrotransposons, respectively.

**Figure S4. SV type composition in the three-caller consensus call sets.** A doughnut plot illustrating the number of three-caller consensus SVs by type detected against GRCh38 (**A**) and T2T-CHM13 (**B**). Stacked bar charts showing the number of three-caller consensus SVs detected per sample in both GRCh38 (**C**) and T2T-CHM13 (**D**). Staked bar plot illustrating the average number of SVs detected per sample for each caller, including the consensus set for GRCh38 (**E**) and T2T-CHM13 (**F**). **(G)** and **(H)** show the size distribution of three-caller consensus SVs for GRCh38 and T2T-CHM13, respectively.

**Figure S5. SV discovery saturation and size distribution patterns.** (**A** and **B**) Cumulative plots showing singleton (red) versus non-singleton (blue) SV accumulation when observing a fixed number of variants as samples are added, demonstrating discovery saturation patterns. (**C** and **D**) represents the number of variants detected per sample, coloured according to allele frequency. A box plot illustrates the size distribution of common and rare SVs (**E** and **F**) GRCh38 and (**G** and **H**) T2T-CHM13.

**Figure S6. Genomic SV burden per sample using GRCh38.** Distribution plots showing the total genomic sequence affected per sample for each caller.

**Figure S7. Genomic SV burden per sample using T2T-CHM13.** Distribution plots showing the total genomic sequence affected per sample for each caller.

**Figure S8. SVAN structural variant classification for the GRCh38 dataset.** Comprehensive breakdown showing classification of 74,364 SVs (76.0% of total) into distinct categories: mobile element insertions (MEIs) including Alu (10,487), L1 (1,632), and SVA (922) elements; variable number tandem repeats (VNTRs) (50,863); duplications (9,245); and other rare classes including NUMTs and processed pseudogenes.

**Figure S9. SVAN structural variant classification for T2T-CHM13 dataset.** Enhanced classification of 136,769 SVs (77.5% of total) showing increased detection of all SV classes compared to GRCh38, particularly VNTRs (112,004) and duplications (14,218), highlighting the superior resolution of complex genomic regions in the complete assembly.

**Figure S10. Boxplots of SV counts in total and separated by type.** The total number of SVs was plotted in a box plot per population from the 1kGP ONT including SVs from MENA individuals, for each genome GRCh38 (**A**) and T2T-CHM13 (**B**). On average, MENA individuals have the lowest Sniffles-called SV count compared to other populations when mapped to GRCh38 (22,448 mean, 22,955 median). However, using the T2T-CHM13, MENA individuals ascended to second place (27,334 mean, 28,321 median), despite contributing the smallest sample size (61 individuals), and surpassed only by African populations (30,384 mean, 30,353 median) (Figure 5) (Table S18). This reversal reflects the ancestry bias of GRCh38, which was eliminated by the gapless T2T-CHM13 assembly. The number of SVs for each population was plotted for each SV type. Insertions (**C** and **D**), Deletions (**E** and **F**) and Duplications (**G** and **H**).

**Figure S11. Pearson correlation coefficients using GRCh38 sniffles-based SVs merged between MEAN and 1kGP ONT.** Pearson correlation coefficients were calculated between MENA and each continental population (AFR, AMR, EAS, EUR, SAS) across all shared SV sites in the merged dataset. SVs were filtered to retain variants with AF $\geq$ 0.01 and AF $\leq$ 0.95 in each compared population, and AF $\geq$ 0.01 and AF $\leq$ 0.90 in MENA, excluding rare variants and near-fixed alleles likely reflecting reference bias. Pearson correlation of SV allele frequencies revealed that MENA populations are most similar to European (r = 0.635) and American (r = 0.624) populations, followed by South Asian (r = 0.613), with lower similarity to East Asian (r = 0.515) and African (r = 0.508) populations. African populations shared the highest absolute number of SV sites with MENA, consistent with the larger reservoir of African structural variation, followed by South Asian populations

**Figure S12. Pearson correlation coefficients using T2T-CHM13 sniffles-based SVs.** The ranking was consistent using T2T-CHM13 (EUR r = 0.549, AMR r = 0.528, SAS r = 0.502, EAS r = 0.424, AFR r = 0.385).

**Figure S13. A boxplot of *Fst* differentiation index for each SV and window size of 1 Mb.**

**Figure S14. *GSTM1* deletion detected when aligning to GRCh38. (**GW) browser screenshots showing a 1.84 kbp deletion affecting *GSTM1* transcripts in samples APR001 (heterozygous) and APR005 (homozygous). Coverage tracks and SV calls demonstrate that the deletion spans multiple exons of this detoxification enzyme gene.

**Figure S15. *CR1* deletions detected using GRCh38 as reference. (**GW) visualisation showing heterozygous deletions in the *CR1* locus for samples APR002 and APR003. APR002 carries a heterozygous 18.5 kb. APR003 displays compound heterozygous deletions with an 18.5 kb deletion (chr1-207543727-DEL->18555) and a partially overlapping 17.1 kb deletion (chr1-207550585-DEL->17089), suggesting complex structural rearrangement at this locus. CR1 is a known structurally discrepant region between GRCh38 and T2T-CHM13, where T2T-CHM13 lacks eight exons (450 amino acids) present in the GRCh38 reference.

**Figure S16. Heterozygous deletion in *KLRC2*. (**GW) visualisation of 15.4 kbp deletions in the *KLRC* region in APR002 and APR008. Coverage plots show heterozygous deletions where one allele resembles the T2T-CHM13 reference (lacking *KLRC2*), and the other resembles GRCh38.

**Figure S17. Enhanced mapping to *MAP2K3* using T2T-CHM13 reference and GRCh38.** Corrected (GW) view showing proper read alignment to *MAP2K3* when using T2T-CHM13, with uniform coverage and no deletion calls, resolving the GRCh38 mapping artefact.

**Figure S18. Deletion in *CD55* detected using GRCh38 as reference.** IGV tracks plotted using (GW) showing a 3.6 kbp deletion in *CD55* with clear breakpoints and reduced coverage. *CD55* encodes a complement regulatory protein involved in malaria susceptibility.

**Figure S19. α-thalassemia 3.8 kb deletion.** Consistent with (– α 3.7 kb single gene deletion), the common α-thalassemia variant, a well-documented deletion with high prevalence in Middle Eastern and Mediterranean populations

**Figure S20. Length distribution of the STR shortest allele.** Box plots comparing short tandem repeat (STR) lengths between MENA samples (orange) and 1000 Genomes ONT data (black) across 68 clinically relevant genes. The length plotted is of the shortest allele length, where the length is pathogenic, indicating an individual with pathogenic expansion on both copies of the gene.

**Figure S21. GRCh38 SVs evolutionary sharing analysis (UpSet plot).** Intersection diagram showing SV sharing patterns between MENA populations, global populations (AFR, EUR, EAS, SAS, AMR), archaic hominins (Neanderthals, Denisovan), and chimpanzee. Bar heights indicate the number of SVs in each intersection category, with population-specific and shared variants clearly delineated.

**Figure S22. T2T-CHM13 SVs evolutionary sharing analysis (UpSet plot).** Enhanced intersection analysis using CHM13-based SVs showing increased detection of population-specific variants and archaic sharing. Demonstrates nearly three-fold increase in MENA-specific variants (18.0% vs 6.1% in GRCh38) and improved resolution of evolutionary relationships between modern and archaic human lineage.

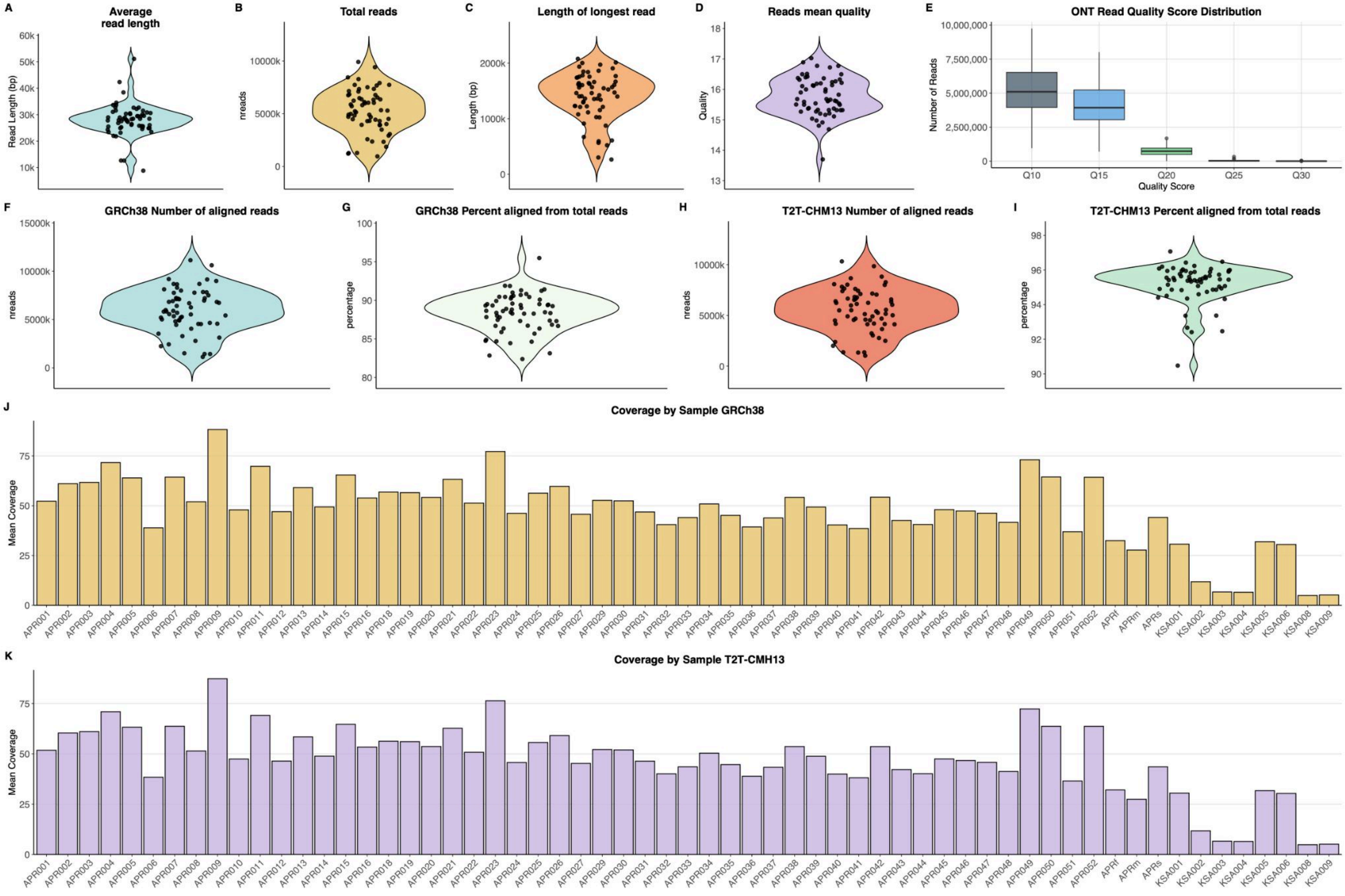
A
Average read length
Read Length (bp)
60k
50k
40k
30k
20k
10k
B
Total reads
nreads
10000k
5000k
0
C
Length of longest read
Length (bp)
2000k
1000k
0
D
Reads mean quality
Quality
18
17
16
15
14
13
E
ONT Read Quality Score Distribution
Number of Reads
10,000,000
7,500,000
5,000,000
2,500,000
0
Q10
Q15
Q20
Q25
Q30
Quality Score
F
GRCh38 Number of aligned reads
nreads
15000k
10000k
5000k
0
G
GRCh38 Percent aligned from total reads
percentage
100
95
90
85
80
H
T2T-CHM13 Number of aligned reads
nreads
10000k
5000k
0
I
T2T-CHM13 Percent aligned from total reads
percentage
98
96
94
92
90
J
Coverage by Sample GRCh38
Mean Coverage
75
50
25
0
APR001 APR002 APR003 APR004 APR005 APR006 APR007 APR008 APR009 APR010 APR011 APR012 APR013 APR014 APR015 APR016 APR018 APR019 APR020 APR021 APR022 APR023 APR024 APR025 APR026 APR027 APR029 APR030 APR031 APR032 APR033 APR034 APR035 APR036 APR037 APR038 APR039 APR040 APR041 APR042 APR043 APR044 APR045 APR046 APR047 APR048 APR049 APR050 APR051 APR052 APRf APRm APRs KSA001 KSA002 KSA003 KSA004 KSA005 KSA006 KSA008 KSA009
K
Coverage by Sample T2T-CMH13
Mean Coverage
75
50
25
0
APR001 APR002 APR003 APR004 APR005 APR006 APR007 APR008 APR009 APR010 APR011 APR012 APR013 APR014 APR015 APR016 APR018 APR019 APR020 APR021 APR022 APR023 APR024 APR025 APR026 APR027 APR029 APR030 APR031 APR032 APR033 APR034 APR035 APR036 APR037 APR038 APR039 APR040 APR041 APR042 APR043 APR044 APR045 APR046 APR047 APR048 APR049 APR050 APR051 APR052 APRf APRm APRs KSA001 KSA002 KSA003 KSA004 KSA005 KSA006 KSA008 KSA009

**Figure S1.**

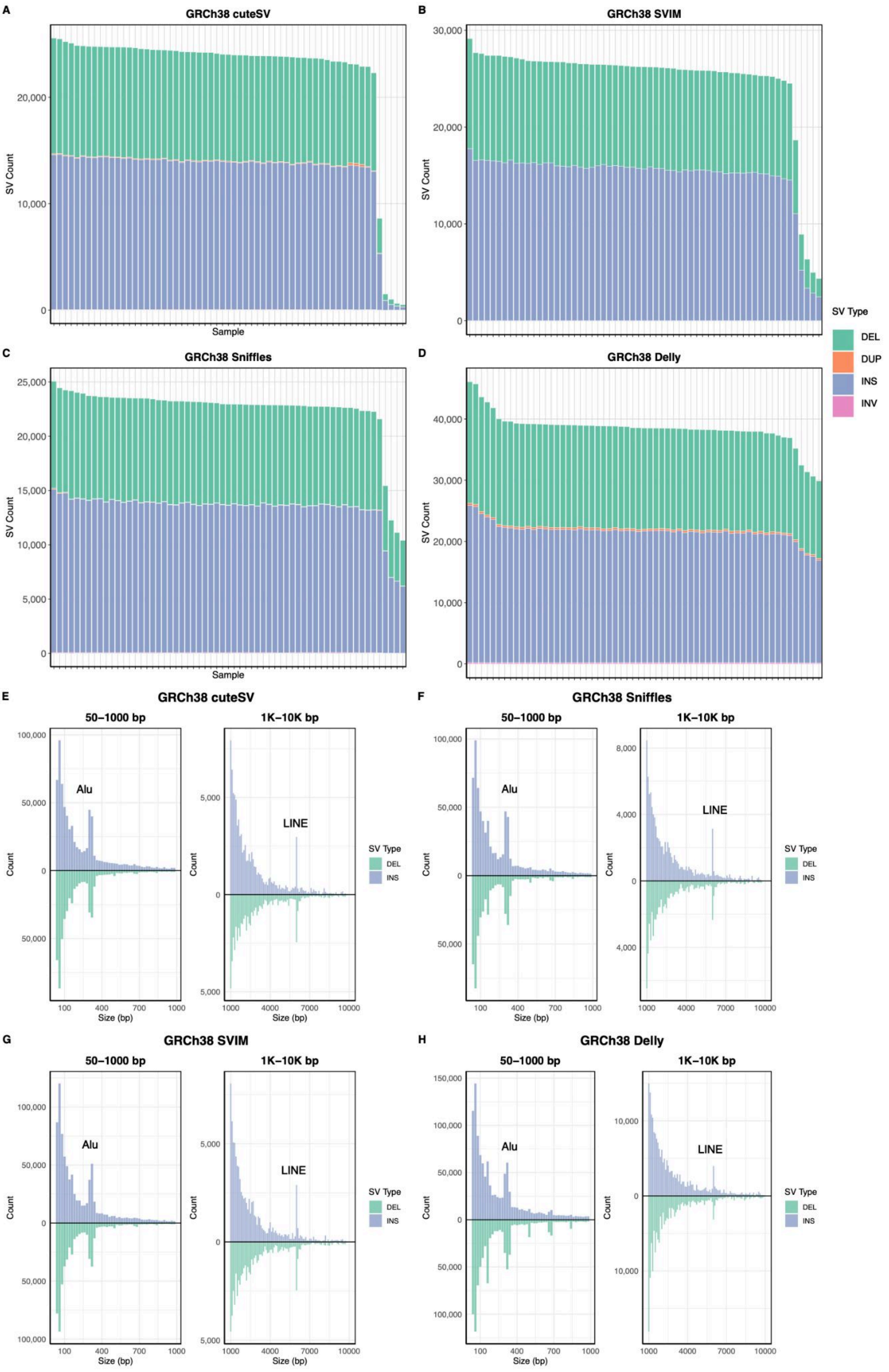
A
GRCh38 cuteSV
B
GRCh38 SVIM
C
GRCh38 Sniffles
D
GRCh38 Delly
SV Count
Sample
SV Type
DEL
DUP
INS
INV
E
GRCh38 cuteSV
F
GRCh38 Sniffles
G
GRCh38 SVIM
H
GRCh38 Delly
50–1000 bp
1K–10K bp
Alu
LINE
Count
Size (bp)
SV Type
DEL
INS

Figure S2.

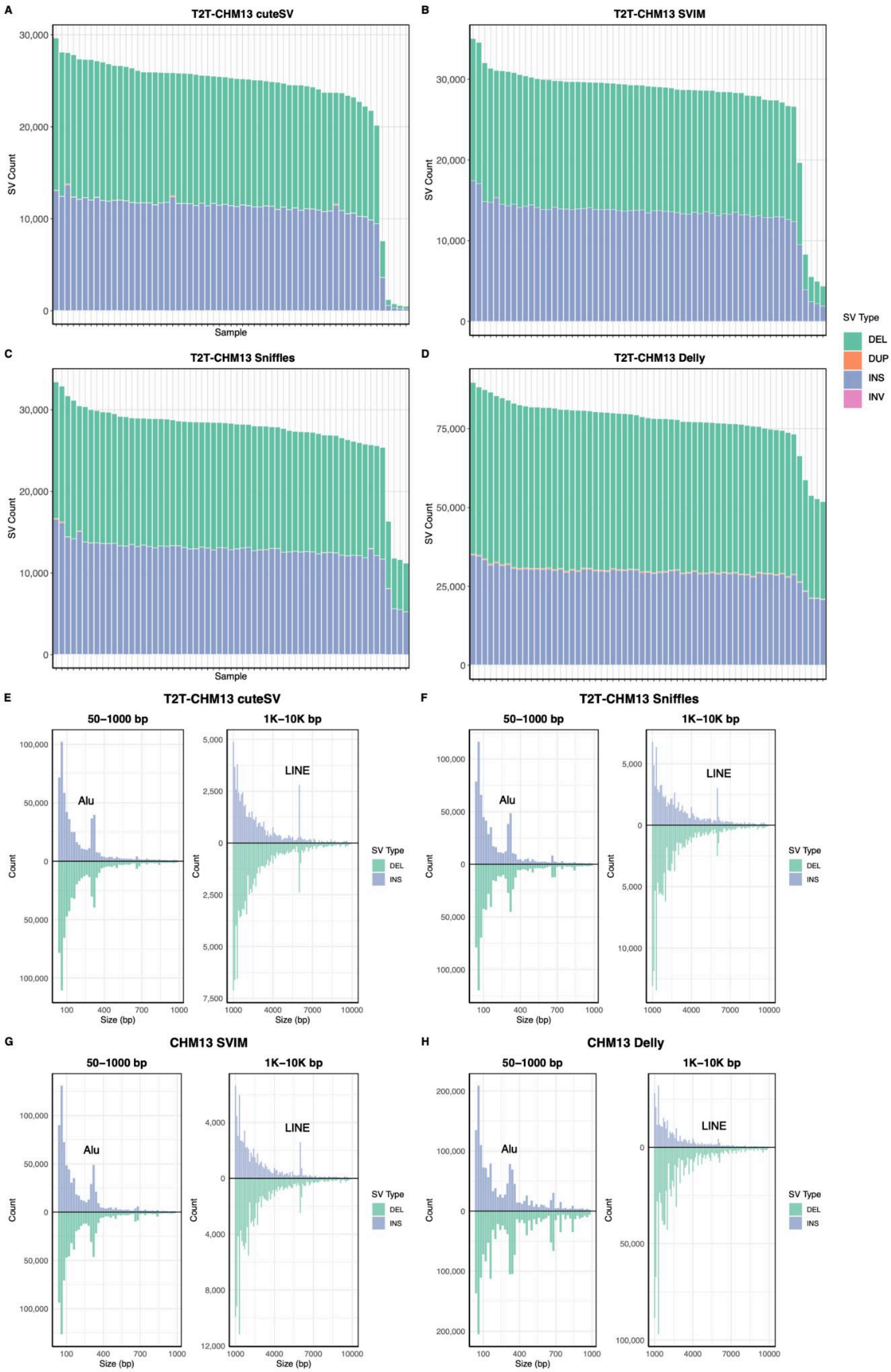
A
T2T-CHM13 cuteSV
SV Count
Sample
B
T2T-CHM13 SVIM
SV Count
C
T2T-CHM13 Sniffles
SV Count
Sample
D
T2T-CHM13 Delly
SV Count
SV Type
DEL
DUP
INS
INV
E
T2T-CHM13 cuteSV
50–1000 bp
1K–10K bp
Alu
LINE
Count
Size (bp)
F
T2T-CHM13 Sniffles
50–1000 bp
1K–10K bp
Alu
LINE
Count
Size (bp)
G
CHM13 SVIM
50–1000 bp
1K–10K bp
Alu
LINE
Count
Size (bp)
H
CHM13 Delly
50–1000 bp
1K–10K bp
Alu
LINE
Count
Size (bp)
SV Type
DEL
INS

**Figure S3.**

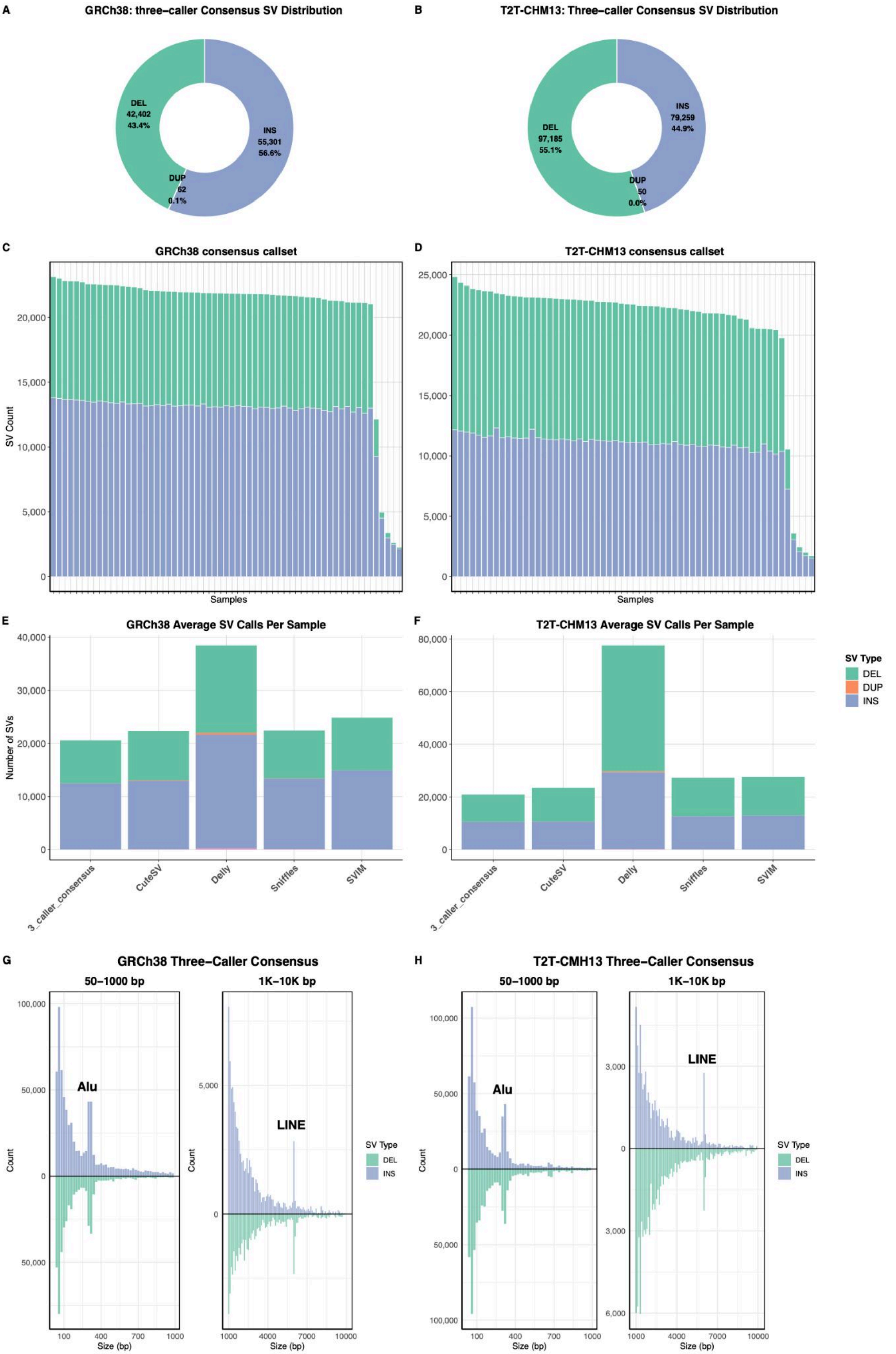
A
GRCh38: three-caller Consensus SV Distribution
DEL
42,402
43.4%
INS
55,301
56.6%
DUP
62
0.1%
B
T2T-CHM13: Three-caller Consensus SV Distribution
INS
79,259
44.9%
DEL
97,185
55.1%
DUP
50
0.0%
C
GRCh38 consensus callset
SV Count
Samples
D
T2T-CHM13 consensus callset
Samples
E
GRCh38 Average SV Calls Per Sample
Number of SVs
3_caller_consensus
CuteSV
Delly
Sniffles
SVIM
F
T2T-CHM13 Average SV Calls Per Sample
SV Type
DEL
DUP
INS
G
GRCh38 Three-Caller Consensus
50–1000 bp
1K–10K bp
Alu
LINE
Count
Size (bp)
H
T2T-CMH13 Three-Caller Consensus
50–1000 bp
1K–10K bp
Alu
LINE

**Figure S4.**

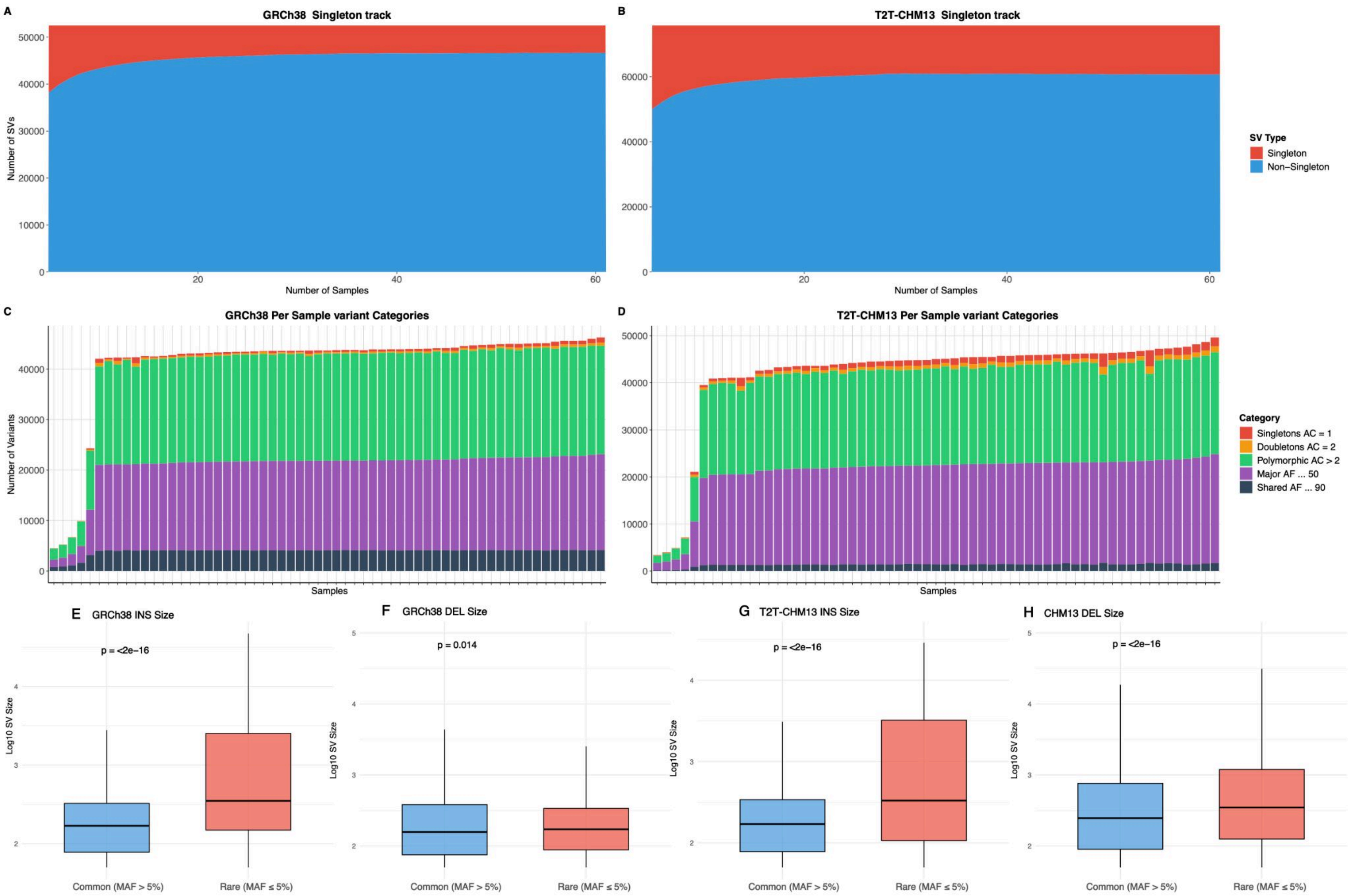

A
GRCh38 Singleton track
Number of SVs
Number of Samples
B
T2T-CHM13 Singleton track
Number of Samples
SV Type
Singleton
Non–Singleton
C
GRCh38 Per Sample variant Categories
Number of Variants
Samples
D
T2T-CHM13 Per Sample variant Categories
Samples
Category
Singletons AC = 1
Doubletons AC = 2
Polymorphic AC > 2
Major AF ... 50
Shared AF ... 90
E GRCh38 INS Size
p = <2e−16
F GRCh38 DEL Size
p = 0.014
G T2T-CHM13 INS Size
p = <2e−16
H CHM13 DEL Size
p = <2e−16
Log10 SV Size
Common (MAF > 5%)
Rare (MAF ≤ 5%)


**Figure S5.**

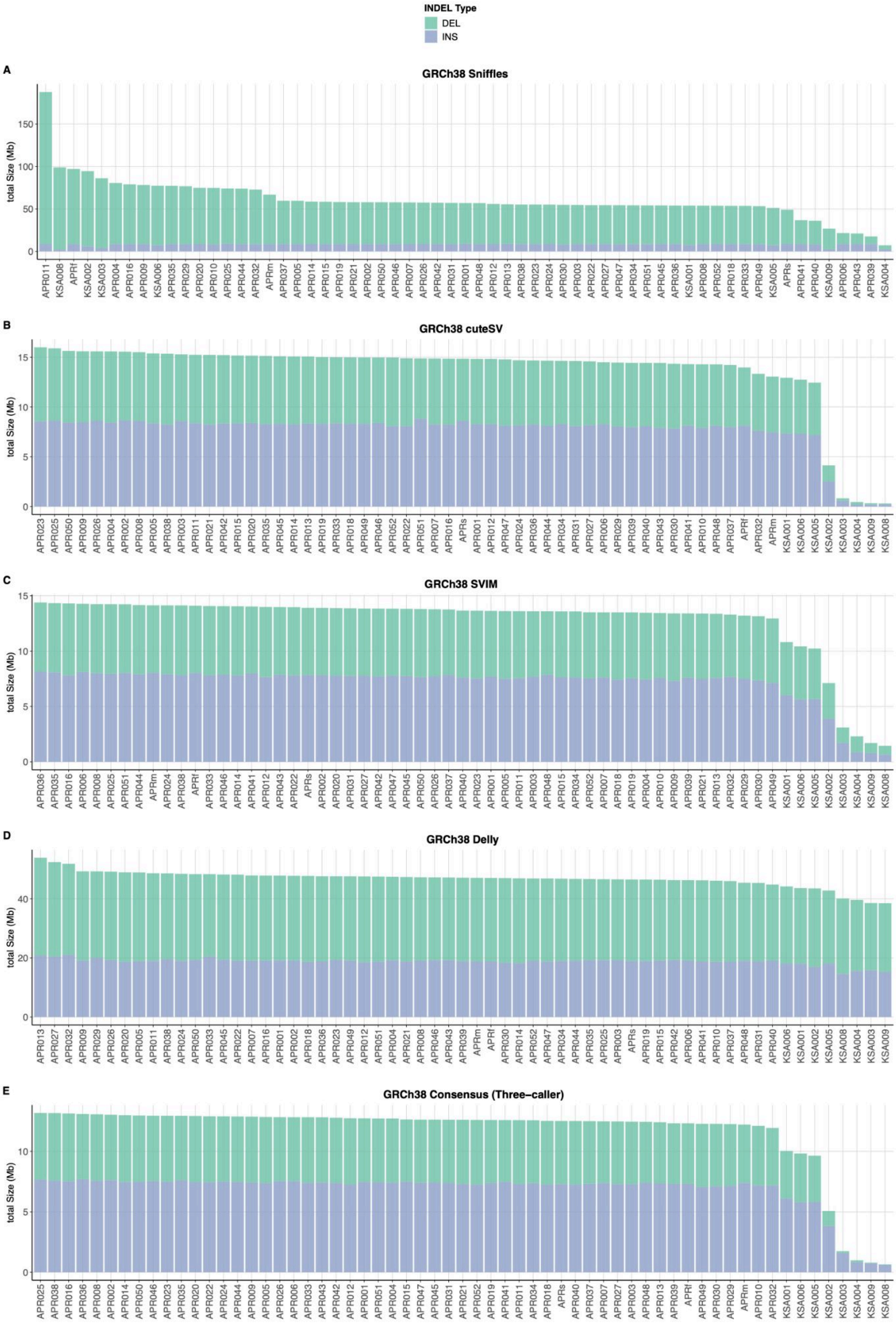
INDEL Type
DEL
INS
A
GRCh38 Sniffles
total Size (Mb)
0
50
100
150
B
GRCh38 cuteSV
total Size (Mb)
0
5
10
15
C
GRCh38 SVIM
total Size (Mb)
0
5
10
15
D
GRCh38 Delly
total Size (Mb)
0
20
40
E
GRCh38 Consensus (Three−caller)
total Size (Mb)
0
5
10

**Figure S6.**

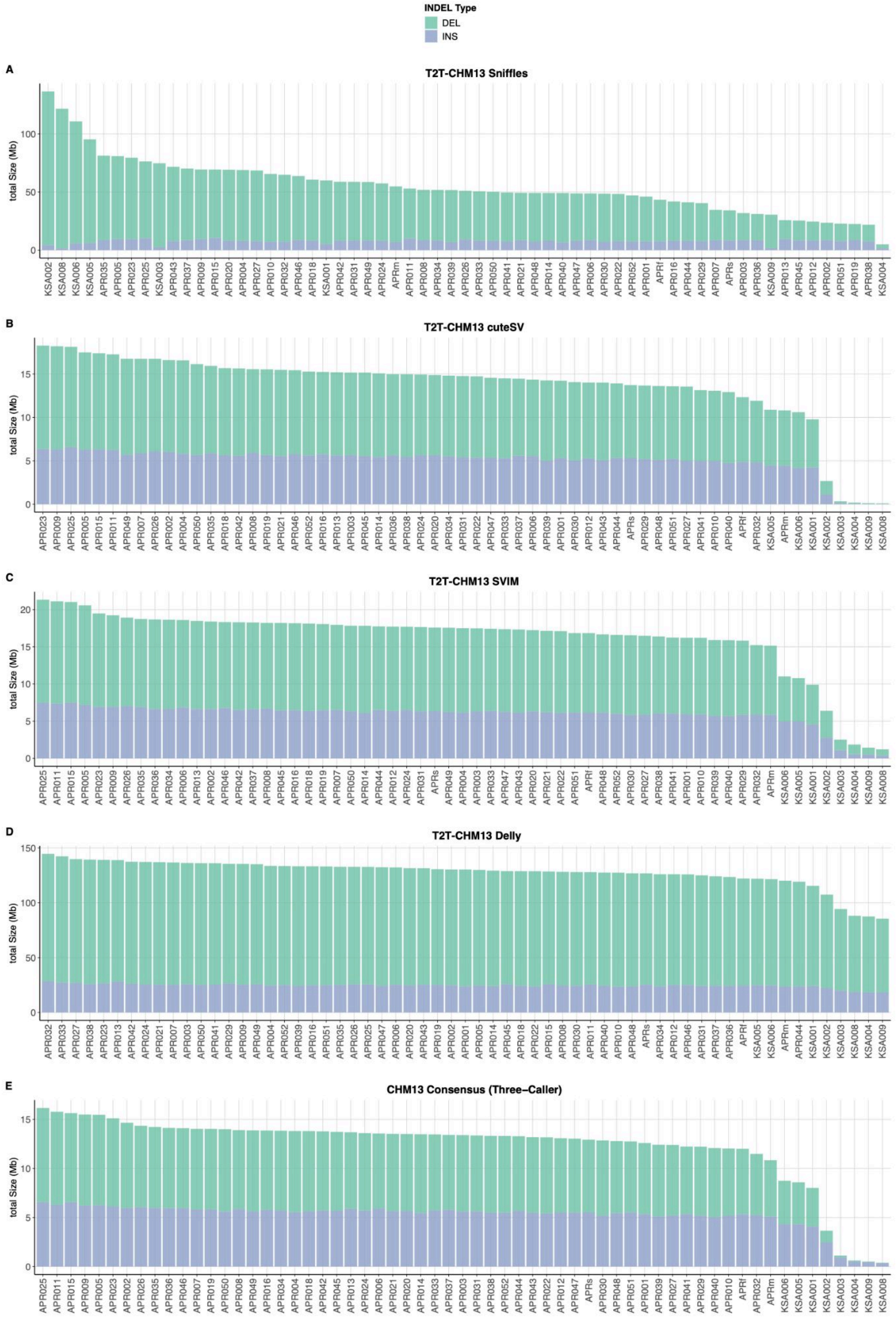
INDEL Type
DEL
INS
A
T2T-CHM13 Sniffles
total Size (Mb)
B
T2T-CHM13 cuteSV
total Size (Mb)
C
T2T-CHM13 SVIM
total Size (Mb)
D
T2T-CHM13 Delly
total Size (Mb)
E
CHM13 Consensus (Three-Caller)
total Size (Mb)

**Figure S7.**

| GRCj38 SV Classes Distribution | | | | | | | | |
|---|---|---|---|---|---|---|---|---|
| SV Class | DEL | INS | Total | % Total | % MAF≤0.05 | % AF≥0.9 | Size Distribution | Mean Length |
| VNTR | 19,424 | 31,477 | 50,901 | 52.04 | | | | 331 |
| Other/Unclassif | 19,442 | 3,911 | 23,415 | 23.94 | | | | 1,171 |
| Alu | 2,366 | 8,121 | 10,487 | 10.72 | | | | 313 |
| Tandem Duplication | 0 | 7,465 | 7,465 | 7.63 | | | | 231 |
| L1 | 372 | 1,260 | 1,632 | 1.67 | | | | 2,496 |
| Duplication Interspersed | 0 | 1,618 | 1,618 | 1.65 | | | | 1,022 |
| Non-canonical MEI | 631 | 551 | 1,182 | 1.21 | | | | 1,955 |
| SVA | 179 | 743 | 922 | 0.94 | | | | 1,588 |
| Inverted duplication | 0 | 81 | 81 | 0.08 | | | | 352 |
| Processed pseudogene | 26 | 41 | 67 | 0.07 | | | | 2,691 |
| NUMT | 0 | 34 | 34 | 0.03 | | | | 640 |
| HERVK | 7 | 24 | 31 | 0.03 | | | | 2,333 |
| Solo-LTR | 1 | 24 | 25 | 0.03 | | | | 822 |
| Complex Duplication | 0 | 23 | 23 | 0.02 | | | | 217 |
| snRNA | 2 | 4 | 6 | 0.01 | | | | 1,034 |

**Figure S8.**

| CHM13 SV Classes Distribution | | | | | | | | |
|---|---|---|---|---|---|---|---|---|
| SV Class | DEL | INS | Total | % Total | % MAF≤0.05 | % AF≥0.9 | Size Distribution | Mean Length |
| VNTR | 63,650 | 48,416 | 112,066 | 63.47 | | | | 773 |
| Other/Unclassif | 33,545 | 6,141 | 39,686 | 22.48 | | | | 927 |
| Tandem Duplication | 0 | 11,324 | 11,324 | 6.41 | | | | 286 |
| Alu | 0 | 8,023 | 8,023 | 4.54 | | | | 305 |
| Duplication Interspersed | 0 | 2,741 | 2,741 | 1.55 | | | | 693 |
| L1 | 0 | 1,239 | 1,239 | 0.70 | | | | 2,590 |
| SVA | 0 | 660 | 660 | 0.37 | | | | 1,754 |
| Non-canonical MEI | 20 | 567 | 587 | 0.33 | | | | 1,914 |
| Inverted duplication | 0 | 86 | 86 | 0.05 | | | | 493 |
| Unannotated | 0 | 0 | 50 | 0.03 | | | | 2,629 |
| NUMT | 0 | 39 | 39 | 0.02 | | | | 1,026 |
| Processed pseudogene | 0 | 36 | 36 | 0.02 | | | | 1,428 |
| HERVK | 0 | 29 | 29 | 0.02 | | | | 2,657 |
| Complex Duplication | 0 | 23 | 23 | 0.01 | | | | 452 |
| Solo-LTR | 0 | 18 | 18 | 0.01 | | | | 879 |
| snRNA | 0 | 3 | 3 | 0.00 | | | | 1,606 |

**Figure S9.**

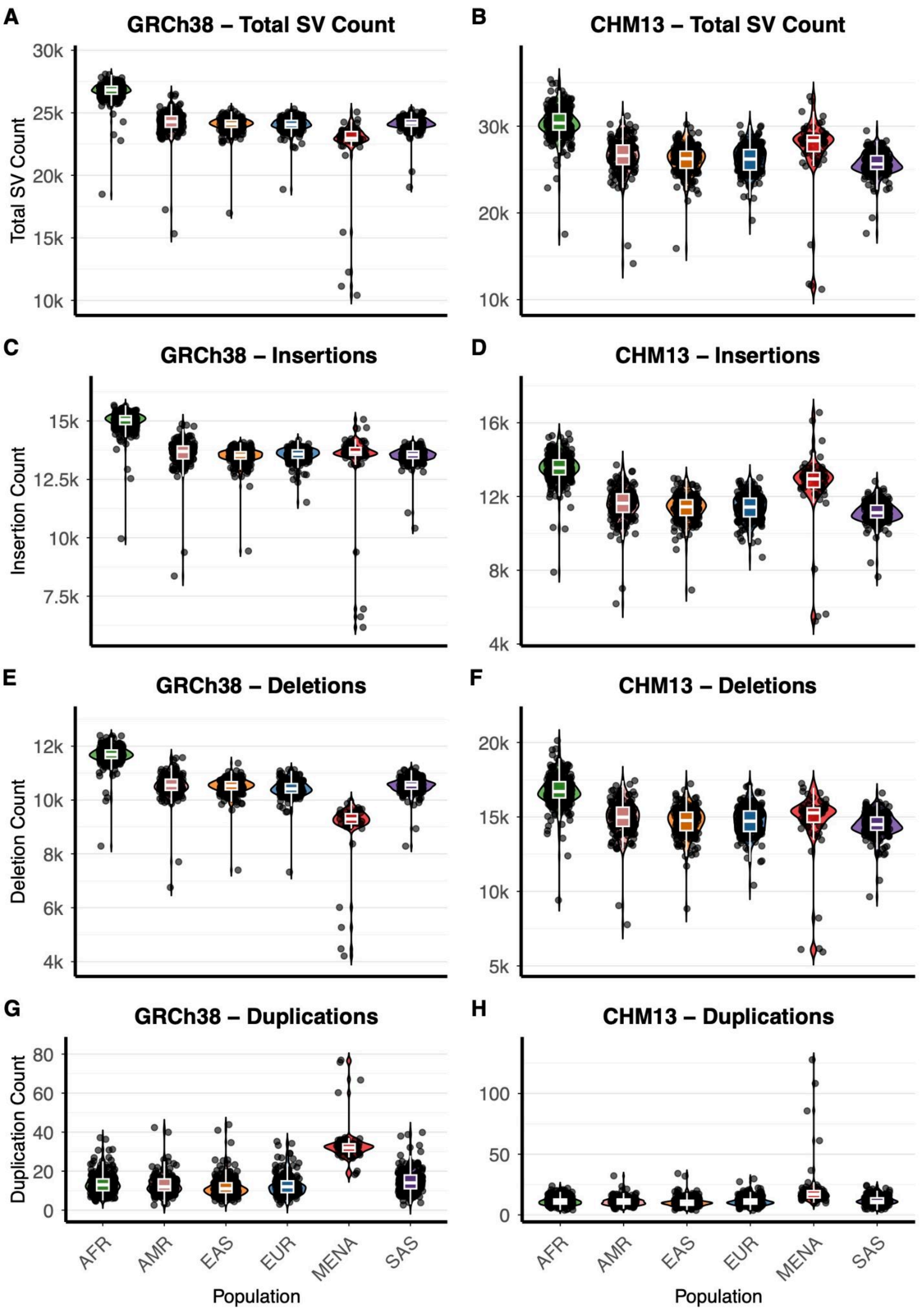
A
GRCh38 – Total SV Count
Total SV Count
30k
25k
20k
15k
10k
B
CHM13 – Total SV Count
30k
20k
10k
C
GRCh38 – Insertions
Insertion Count
15k
12.5k
10k
7.5k
D
CHM13 – Insertions
16k
12k
8k
4k
E
GRCh38 – Deletions
Deletion Count
12k
10k
8k
6k
4k
F
CHM13 – Deletions
20k
15k
10k
5k
G
GRCh38 – Duplications
Duplication Count
80
60
40
20
0
H
CHM13 – Duplications
100
50
0
AFR
AMR
EAS
EUR
MENA
SAS
Population

**Figure S10.**

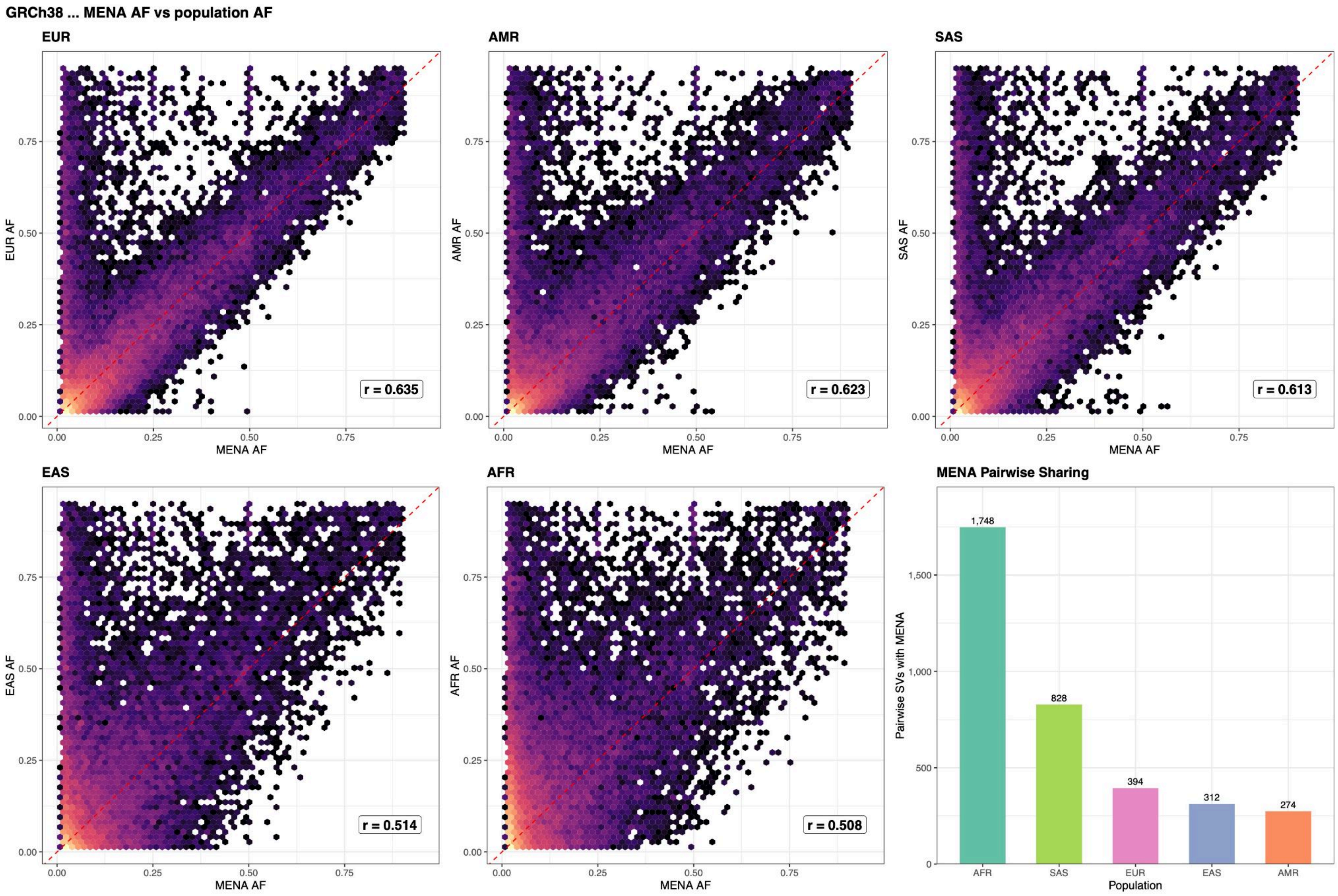
GRCh38 ... MENA AF vs population AF
EUR
EUR AF
MENA AF
r = 0.635
AMR
AMR AF
MENA AF
r = 0.623
SAS
SAS AF
MENA AF
r = 0.613
EAS
EAS AF
MENA AF
r = 0.514
AFR
AFR AF
MENA AF
r = 0.508
MENA Pairwise Sharing
Pairwise SVs with MENA
Population
1,748
828
394
312
274
AFR
SAS
EUR
EAS
AMR

**Figure S11.**

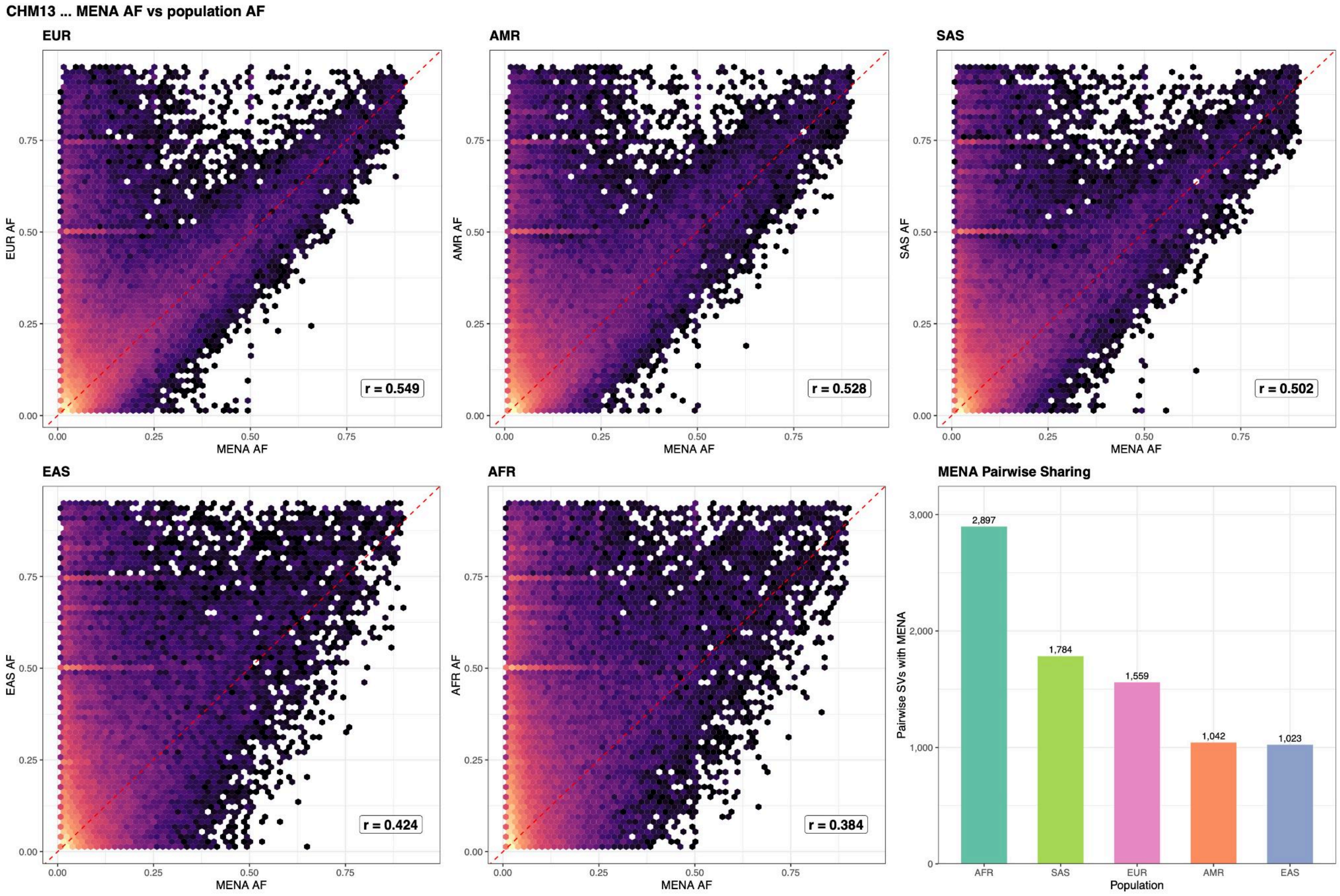
CHM13 ... MENA AF vs population AF
EUR
AMR
SAS
EAS
AFR
MENA Pairwise Sharing
EUR AF
AMR AF
SAS AF
EAS AF
AFR AF
MENA AF
0.00
0.25
0.50
0.75
r = 0.549
r = 0.528
r = 0.502
r = 0.424
r = 0.384
Pairwise SVs with MENA
0
1,000
2,000
3,000
2,897
1,784
1,559
1,042
1,023
AFR
SAS
EUR
AMR
EAS
Population

**Figure S12.**

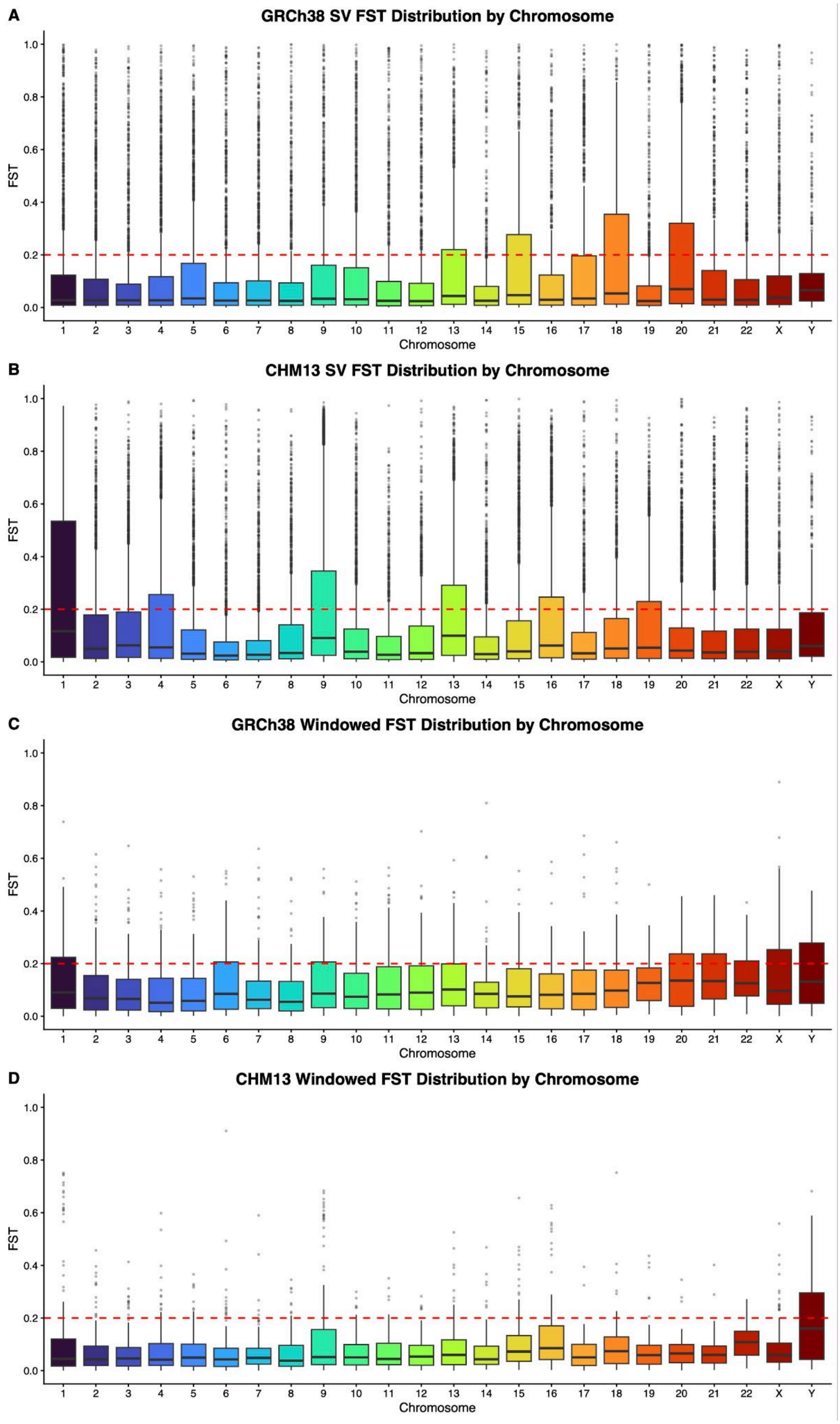
A
GRCh38 SV FST Distribution by Chromosome
B
CHM13 SV FST Distribution by Chromosome
C
GRCh38 Windowed FST Distribution by Chromosome
D
CHM13 Windowed FST Distribution by Chromosome
FST
0.0
0.2
0.4
0.6
0.8
1.0
Chromosome
1 2 3 4 5 6 7 8 9 10 11 12 13 14 15 16 17 18 19 20 21 22 X Y

**Figure S13.**

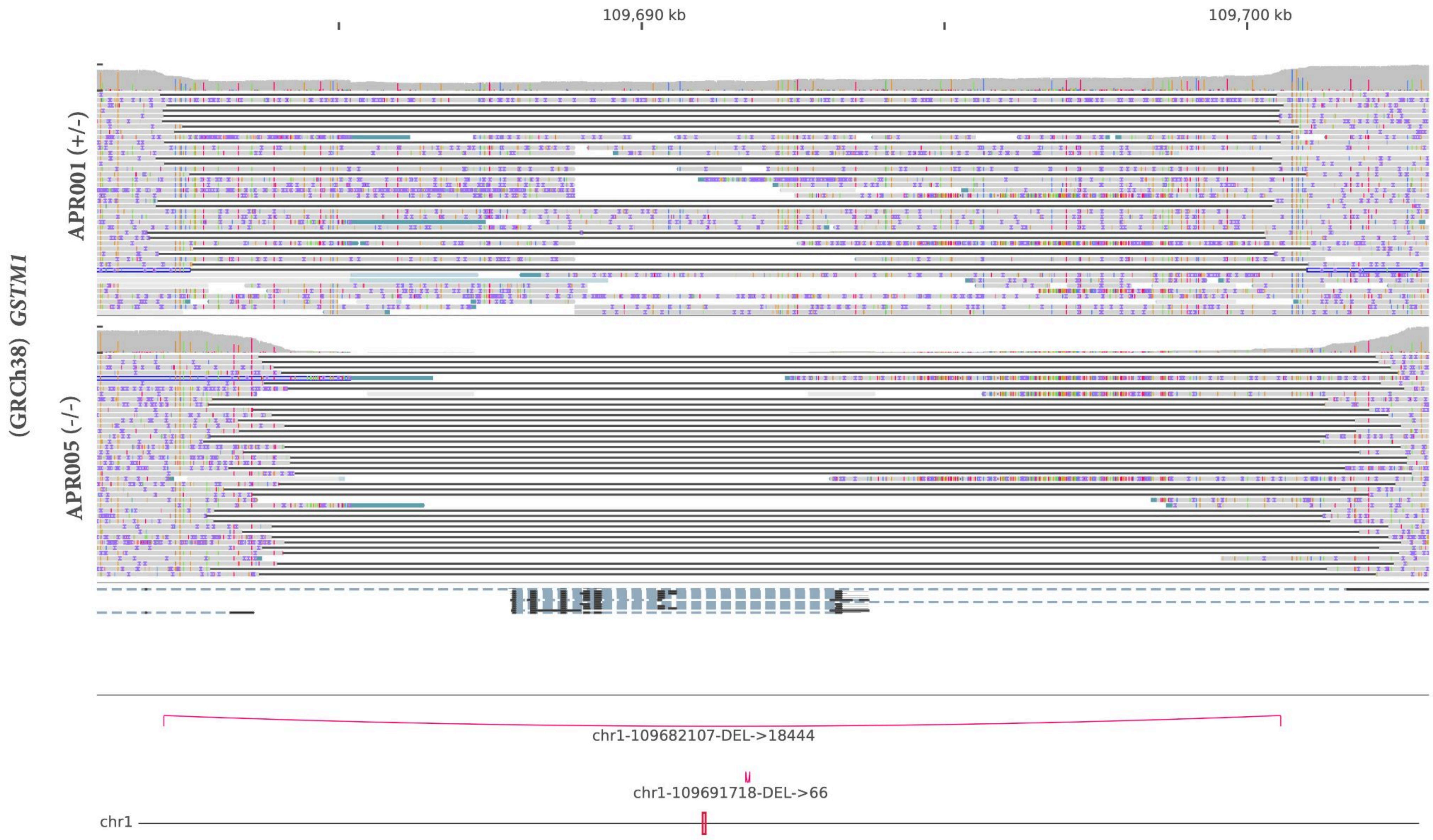

109,690 kb
109,700 kb
APR001 (+/-)
APR005 (-/-)
(GRCh38) GSTM1
chr1-109682107-DEL->18444
chr1-109691718-DEL->66
chr1


**Figure S14.**

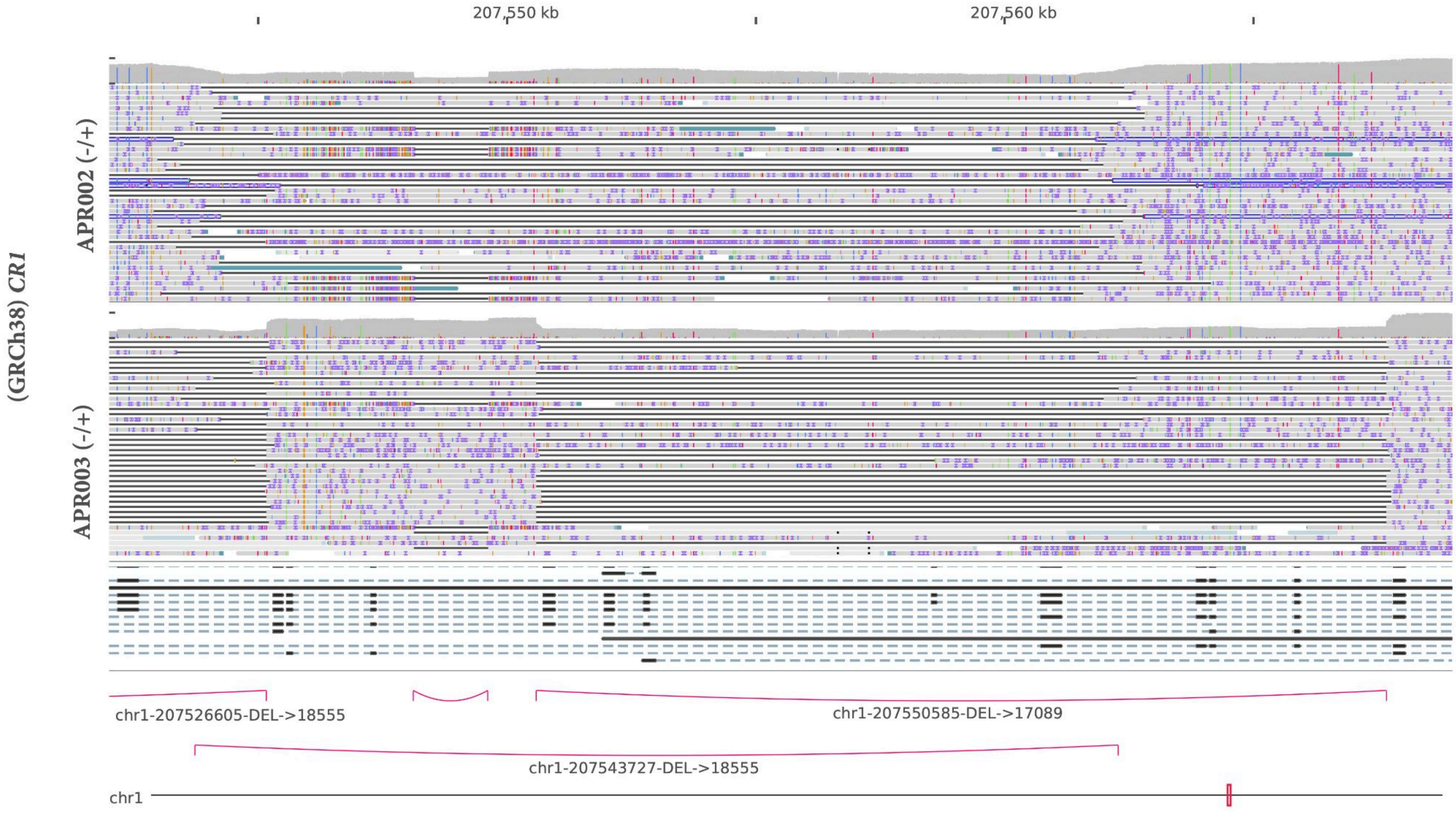
207,550 kb
207,560 kb
(GRCh38) CR1
APR002 (-/+)
APR003 (-/+)
chr1-207526605-DEL->18555
chr1-207550585-DEL->17089
chr1-207543727-DEL->18555
chr1

**Figure S15.**

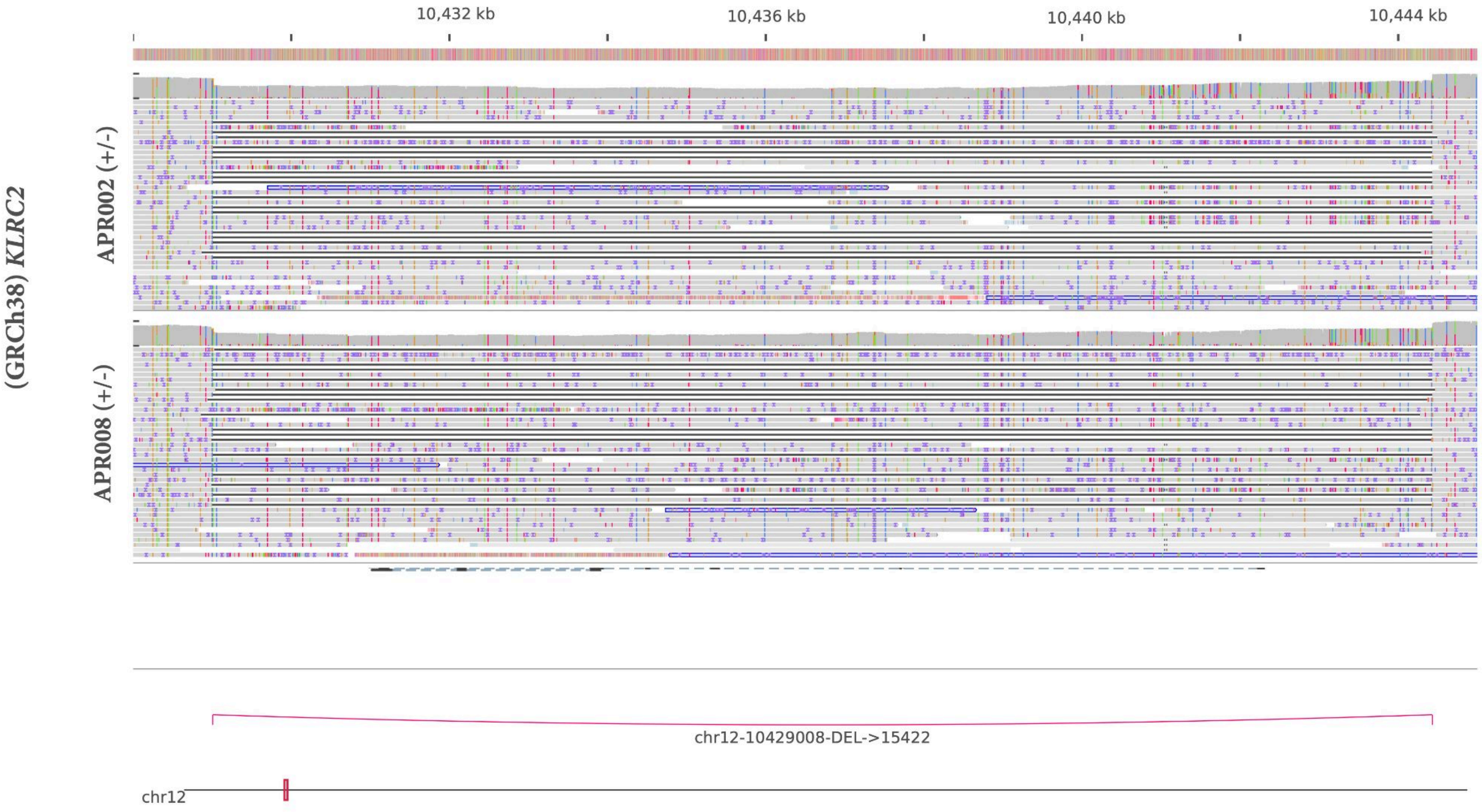
(GRCh38) KLRC2
APR002 (+/-)
APR008 (+/-)
10,432 kb
10,436 kb
10,440 kb
10,444 kb
chr12-10429008-DEL->15422
chr12

**Figure S16.**

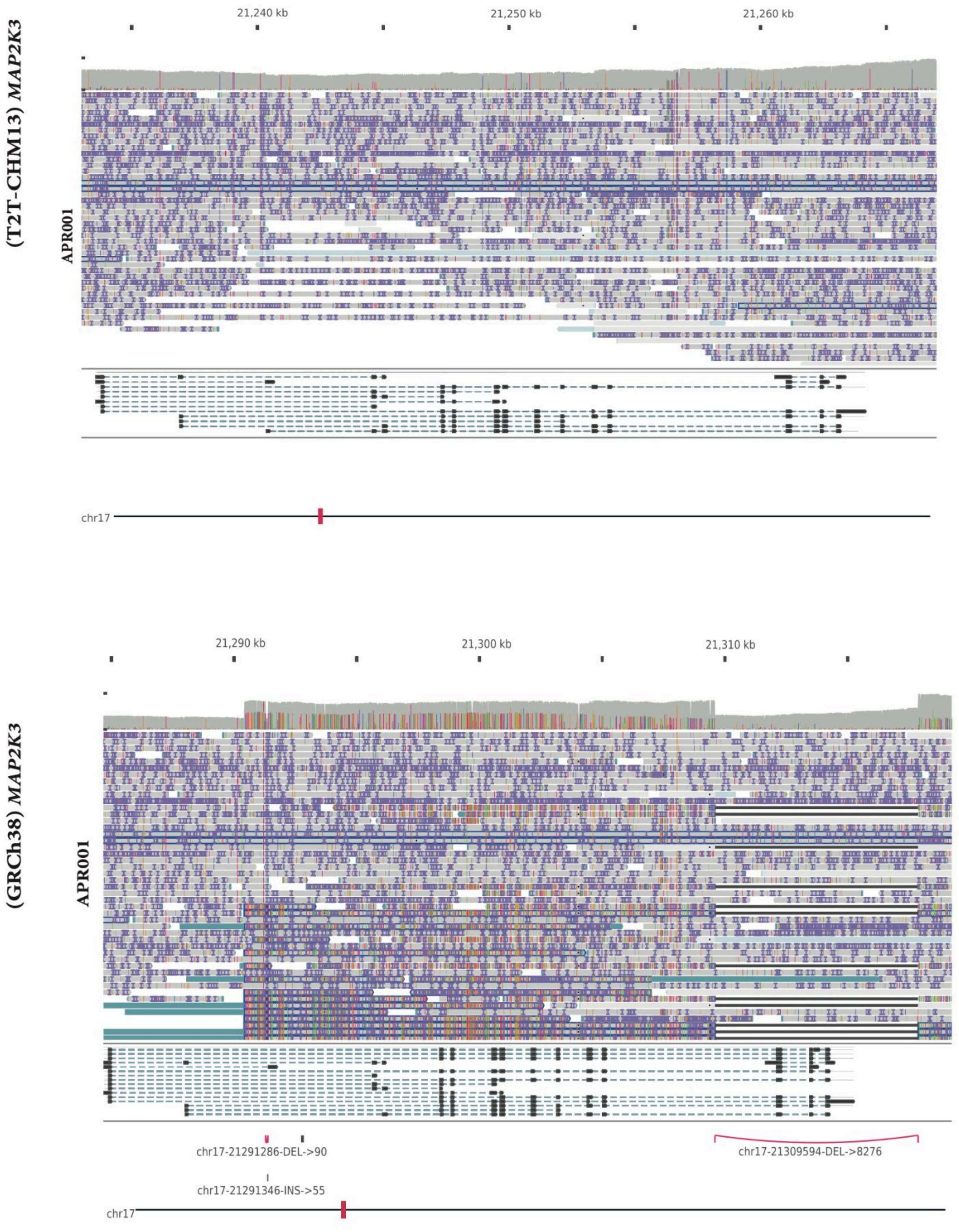

(T2T-CHM13) MAP2K3
APR001
21,240 kb
21,250 kb
21,260 kb
chr17
(GRCh38) MAP2K3
APR001
21,290 kb
21,300 kb
21,310 kb
chr17-21291286-DEL->90
chr17-21309594-DEL->8276
chr17-21291346-INS->55
chr17


**Figure S17.**

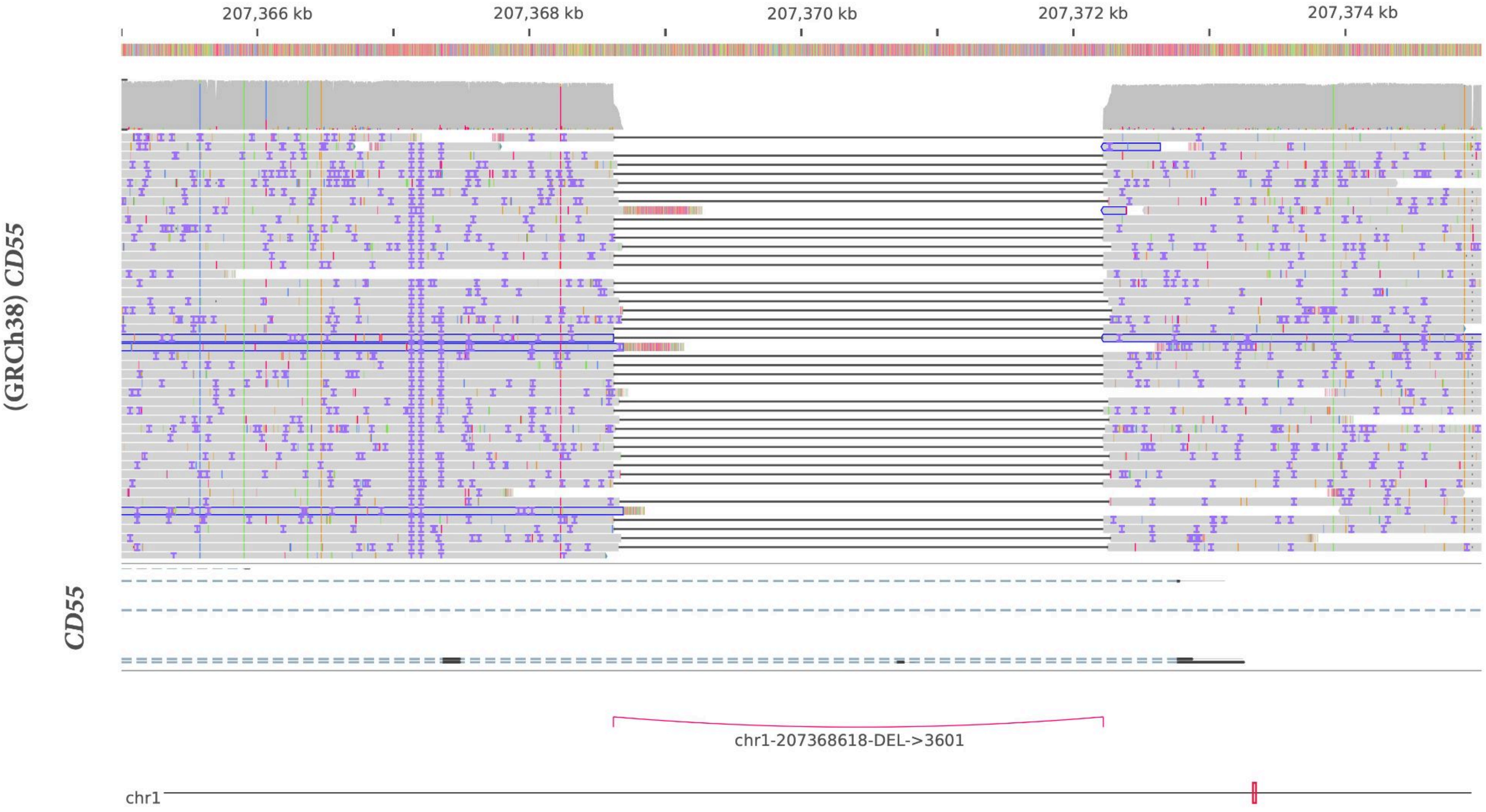

207,366 kb
207,368 kb
207,370 kb
207,372 kb
207,374 kb
(GRCh38) CD55
CD55
chr1-207368618-DEL->3601
chr1


**Figure S18.**

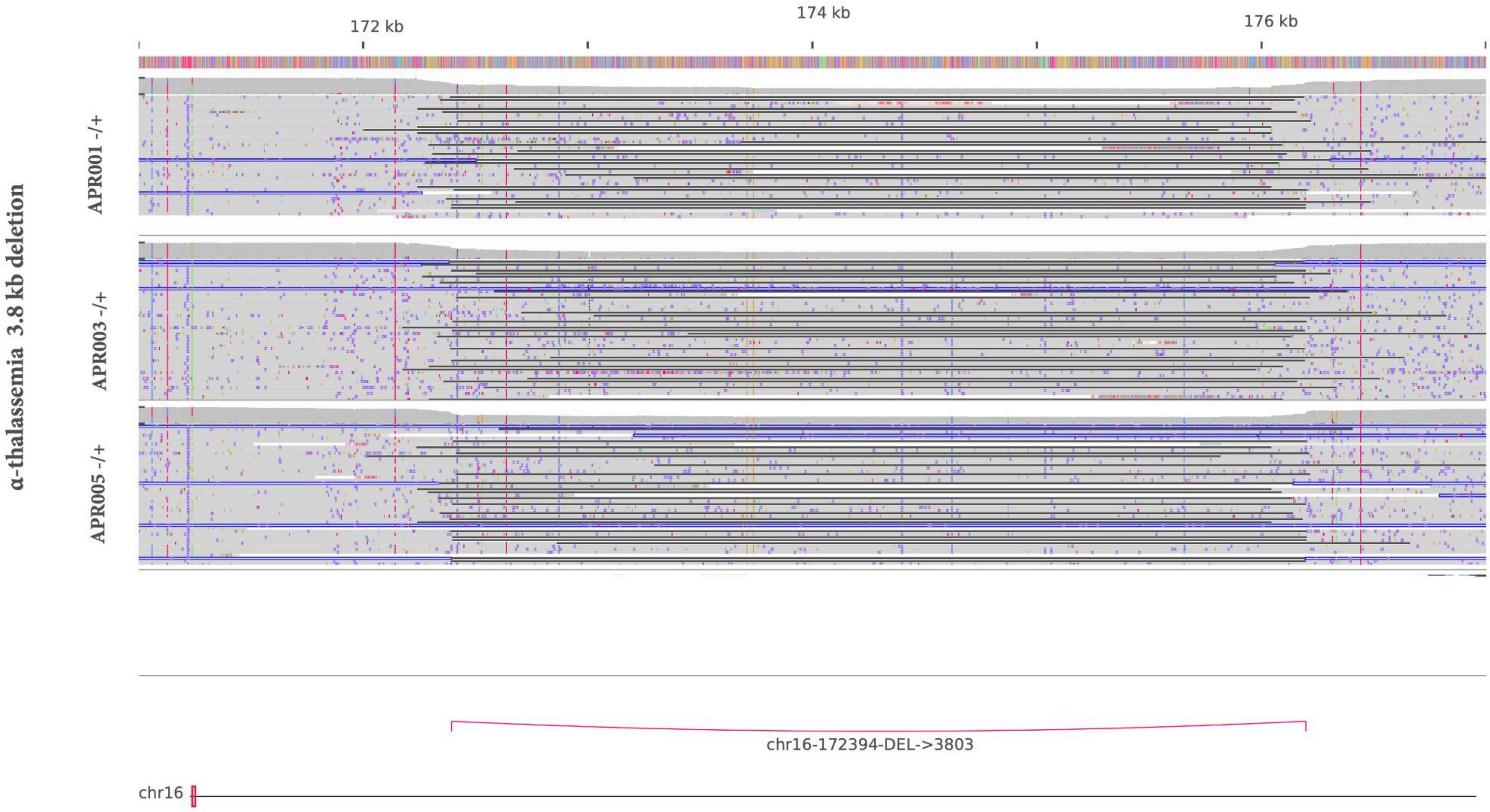
α-thalassemia 3.8 kb deletion
172 kb
174 kb
176 kb
APR001 -/+
APR003 -/+
APR005 -/+
chr16-172394-DEL->3803
chr16

**Figure S19.**

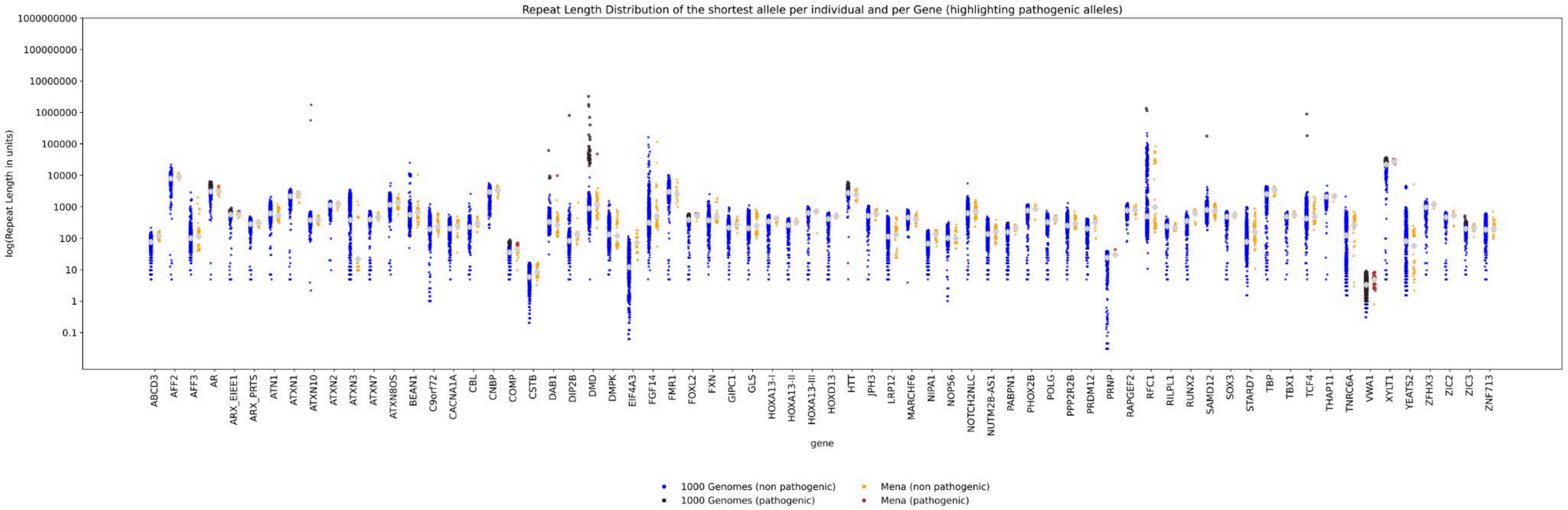

Repeat Length Distribution of the shortest allele per individual and per Gene (highlighting pathogenic alleles)
log(Repeat Length in units)
1000000000
100000000
10000000
1000000
100000
10000
1000
100
10
1
0.1
gene
ABCD3
AFF2
AFF3
AR
ARX_EIEE1
ARX_PRTS
ATN1
ATXN1
ATXN10
ATXN2
ATXN3
ATXN7
ATXN8OS
BEAN1
C9orf72
CACNA1A
CBL
CNBP
COMP
CSTB
DAB1
DIP2B
DMD
DMPK
EIF4A3
FGF14
FMR1
FOXL2
FXN
GIPC1
GLS
HOXA13-I
HOXA13-II
HOXA13-III
HOXD13
HTT
JPH3
LRP12
MARCHF6
NIPA1
NOP56
NOTCH2NLC
NUTM2B-AS1
PABPN1
PHOX2B
POLG
PPP2R2B
PRDM12
PRNP
RAPGEF2
RFC1
RILPL1
RUNX2
SAMD12
SOX3
STARD7
TBP
TBX1
TCF4
THAP11
TNRC6A
VWA1
XYLT1
YEATS2
ZFHX3
ZIC2
ZIC3
ZNF713
1000 Genomes (non pathogenic)
1000 Genomes (pathogenic)
Mena (non pathogenic)
Mena (pathogenic)


**Figure S20.**

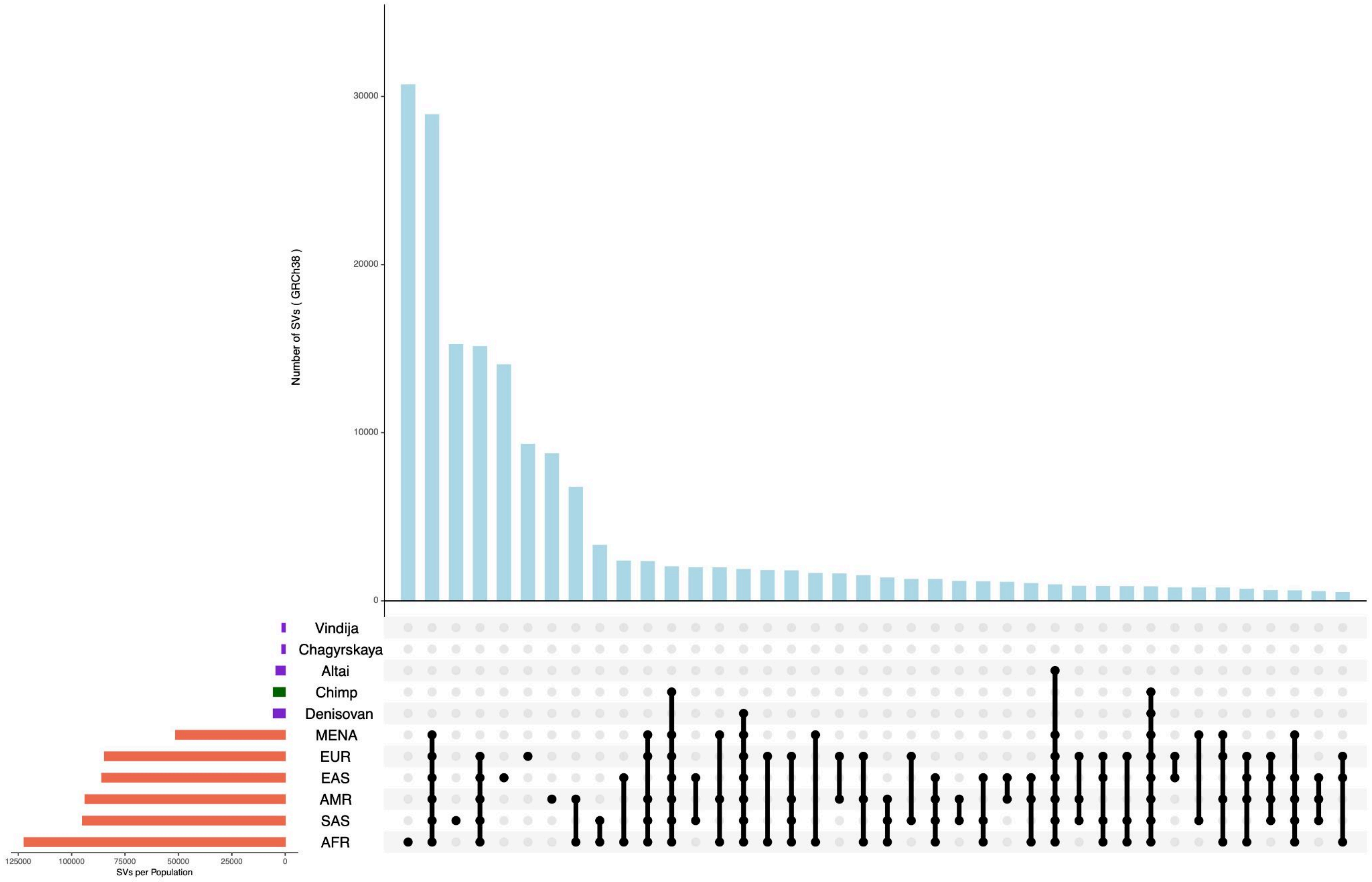
30000
20000
10000
0
Number of SVs ( GRCh38 )
Vindija
Chagyrskaya
Altai
Chimp
Denisovan
MENA
EUR
EAS
AMR
SAS
AFR
125000
100000
75000
50000
25000
0
SVs per Population

**Figure S21.**

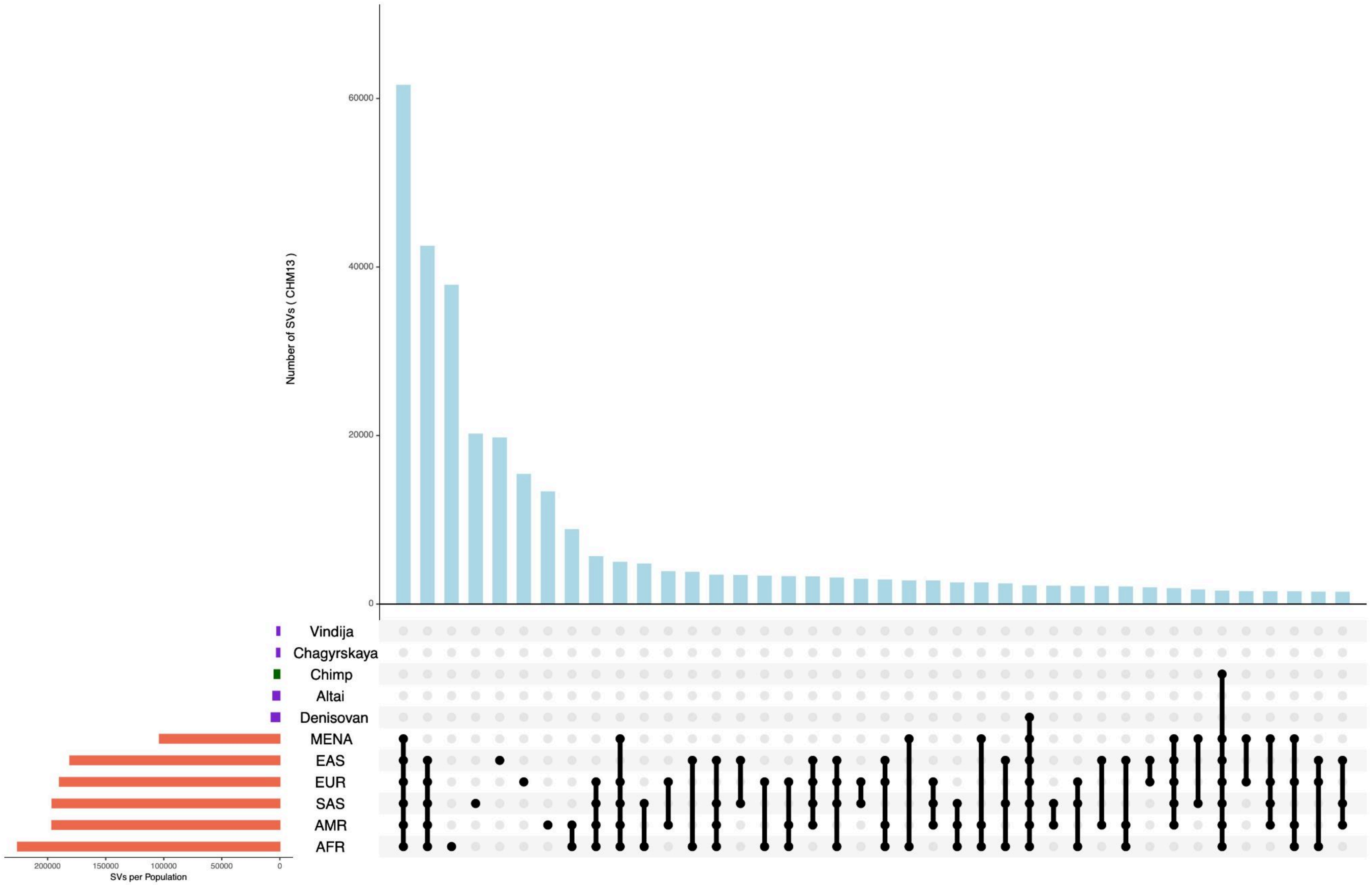

Number of SVs ( CHM13 )
60000
40000
20000
0
Vindija
Chagyrskaya
Chimp
Altai
Denisovan
MENA
EAS
EUR
SAS
AMR
AFR
200000
150000
100000
50000
0
SVs per Population


**Figure S22.**
